# AI-Enabled Echocardiography Identifies Prominent Epicardial Adipose Tissue Associated With an Adverse Cardiometabolic Profile

**DOI:** 10.64898/2026.07.22.26358690

**Authors:** Arya Aminorroaya, Andreas Coppi, Robert L. McNamara, Joao A.C. Lima, Harlan M. Krumholz, Charalambos Antoniades, Rohan Khera, Evangelos K. Oikonomou

**Affiliations:** Section of Cardiovascular Medicine, Department of Internal Medicine, Yale School of Medicine, New Haven, CT, USA; Cardiovascular Data Science (CarDS) Lab, Yale School of Medicine, New Haven, CT, USA; Division of Cardiology, Department of Medicine, Johns Hopkins University School of Medicine, Baltimore, MD, USA; Department of Health Policy and Management, Yale School of Public Health, New Haven, CT, USA; Center for Outcomes Research and Evaluation, Yale-New Haven Hospital, New Haven, CT, USA; Division of Cardiovascular Medicine, Radcliffe Department of Medicine, University of Oxford, Oxford, United Kingdom; Section of Health Informatics, Department of Biostatistics, Yale School of Public Health, New Haven, CT, USA; Department of Biomedical Informatics and Data Science, Yale School of Medicine, New Haven, CT, USA

**Keywords:** artificial intelligence, echocardiography, epicardial adipose tissue, cardiovascular-kidney-metabolic syndrome

## Abstract

**Background:** Excess epicardial adipose tissue (EAT) is associated with cardiovascular-kidney-metabolic (CKM) dysfunction, but its assessment has traditionally required advanced imaging.

**Objectives:** We investigated whether AI-enabled echocardiography could characterize epicardial adiposity and identify residual cardiometabolic risk beyond body mass index (BMI).

**Methods:** We developed *PanAdipo*, a video-based deep learning model in 1,114,441 videos from 28,797 studies across the Yale-New Haven Health System (YNHHS; 2016-2022), using report-derived expert reader labels of prominent EAT (2.3% of studies). *PanAdipo* was evaluated in five cohorts: a *temporally* distinct YNHHS transthoracic echocardiography (TTE) set (n=4,588 patients contributing 5,130 studies), an emergency department point-of-care ultrasound (POCUS) cohort (n=10,957), the geographically distinct MIMIC-IV dataset (n=4,549), the community-based Multi-Ethnic Study of Atherosclerosis (MESA Exam 6, n=2,740), and in held-out YNHHS patients with paired coronary CT angiography within 12 months (n=5,167). Analyses examined discrimination of the report label and of CT-defined high indexed EAT volume, correlation with paired cardiac CT phenotypes, and BMI-adjusted associations with cardiometabolic biomarkers and newly documented metabolic disease.

**Results:** In the held-out health system test set, PanAdipo identified prominent EAT documented in clinical reports with an area under the receiver operating characteristic curve (AUROC) of 0.86 (95% CI, 0.82-0.91) across all views and of up to 0.93 when combining apical 4-chamber and parasternal views. Against paired CT-derived measurements, PanAdipo discriminated age- and sex-specific high EAT (EAT/heart volume ≥95th percentile) with AUROCs of 0.91 to 0.95 across age-sex groups and correlated more strongly with indexed EAT volume (ρ=0.76) than with BMI (ρ=0.41, all P<0.001); PanAdipo alone explained 53% of indexed EAT volume variance, versus 23% for age, sex, and BMI combined. In MESA, higher scores were associated with an adverse metabolic profile (higher HOMA-IR [β=0.17 per SD; 95% CI, 0.11-0.22]) after adjustment for age, sex, and BMI. In health system TTE and POCUS cohorts, higher scores were associated with newly documented MASLD/MASH (BMI-adjusted HR_pooled_ 1.31 [1.03-1.67] per SD of log[PanAdipo]) and hyperlipidemia (1.11 [1.04-1.18]).

**Conclusions:** AI-enabled echocardiography provides a scalable digital biomarker that characterizes prominent epicardial adiposity and is associated with an adverse cardiometabolic profile, highlighting a role for echocardiography in metabolic risk stratification.

**CONDENSED ABSTRACT:** Epicardial adipose tissue (EAT) is linked to cardiometabolic risk, but its assessment has traditionally required advanced imaging. We developed PanAdipo, a deep learning model that derives a digital biomarker of EAT from routine echocardiography. PanAdipo tracks CT-defined epicardial adiposity (ρ=0.76; AUROC 0.91-0.95 for age-/sex-specific high indexed-EAT), yet correlates modestly with body mass index (BMI; ρ=0.19-0.41). After BMI adjustment, higher scores were associated with insulin resistance in a community-based cohort and newly documented metabolic disease across health systems. AI-enabled echocardiography may provide a scalable window into epicardial adiposity, with potential applications for enriching trial populations and monitoring response to cardiometabolic therapies.

## INTRODUCTION

Cardiovascular-kidney-metabolic (CKM) syndrome, as recently defined by the American Heart Association (AHA), frames the pathophysiologic continuum that links excess and dysfunctional adiposity to type 2 diabetes (T2D), chronic kidney disease, atherosclerotic cardiovascular disease, and heart failure.^1^ Within this construct, dysfunctional adipose tissue is the proximate driver of cardiometabolic risk,^2^ and CKM staging explicitly identifies adiposity as a target for early intervention.^1–3^ However, body mass index (BMI), the dominant clinical descriptor of adiposity, only partially reflects the type, location, and biological behavior of the fat that determines risk.^2,4^ Visceral and ectopic depots, in particular epicardial adipose tissue (EAT), are more closely linked to insulin resistance and dyslipidemia than total body weight, even across the normal BMI range.^4–8^

EAT is uniquely positioned over the myocardium and coronary arteries, shares its microcirculation with the heart, and exchanges paracrine and vasocrine signals with the adjacent myocardium.^8–10^ Beyond its volume, the composition of EAT is also informative. Radiomic phenotyping of EAT on cardiac computed tomography (CT) encodes early myocardial remodeling and predicts future heart failure before clinical onset.^11,12^ EAT is also a clinically actionable target, responsive to weight loss, glucagon-like peptide-1 receptor agonists (GLP-1 RAs), and sodium-glucose cotransporter-2 (SGLT2) inhibitors, each of which carries CKM benefits beyond glycemic control.^13,14^ Despite this biological and therapeutic relevance, scalable phenotyping of EAT remains confined to dedicated cardiac CT or magnetic resonance imaging,^5,13^ which restricts its use for population-level prevention and trial design.

Echocardiography is one of the most widely performed cardiac imaging modalities and directly captures the epicardial fat compartment.^15,16^ Prior work has established the feasibility of manually measuring EAT thickness from the parasternal long-axis (PLAX) view;^16,17^ yet manual quantification remains operator-dependent, view-restricted, and difficult to deploy at scale. Recent advances in foundation models for echocardiography enable view-agnostic, multi-task interpretation from both protocolized transthoracic echocardiography (TTE) and point-of-care ultrasound (POCUS) acquisitions.^18,19^ However, existing tools have focused on ventricular function, chamber geometry, and valvular phenotypes, and do not capture epicardial adiposity.

Here, we present *PanAdipo*, a video-based deep learning model that derives a digital biomarker of epicardial fat from standard and portable echocardiographic acquisitions. Across temporally and geographically distinct health system and community-based cohorts, we evaluated whether this AI-derived phenotype captures information beyond existing echocardiographic measurements, corresponds to distinct adiposity phenotypes on paired cross-sectional imaging, and is associated with both cardiometabolic biomarkers and subsequently documented metabolic disease. This work seeks to open a new avenue for echocardiography, extending its role beyond the characterization of cardiac structure and function to metabolic risk stratification using images already acquired as part of routine clinical care.

## METHODS

### Study overview and ethics

This was a multi-cohort, retrospective study of AI-enabled echocardiographic phenotyping of prominent epicardial adiposity. The study was approved by the Yale Institutional Review Board with a waiver of informed consent (Protocol #2000040942). Analyses of de-identified data from Medical Information Mart for Intensive Care, version IV (MIMIC-IV and MIMIC-IV-Echo), accessed through PhysioNet-credentialed access,^20,21^ and the Multi-Ethnic Study of Atherosclerosis (MESA), accessed through NHLBI BioLINCC and BioData Catalyst, were performed under approved data use agreements.^22^

### Development cohort and label definition

We developed PanAdipo using 1,114,441 videos from 28,797 random TTEs in 21,479 unique patients performed across Yale-New Haven Health System (YNHHS) hospitals and outpatient clinics between January 1, 2016, and June 30, 2022, building on the curated multi-view dataset assembled for PanEcho, as described below.^18^ The training label was an expert reader-assigned label for a prominent epicardial fat depot, recorded by board-certified echocardiographers during routine clinical reporting in the structured findings and conclusions fields of the final report. We extracted the label at scale using a deterministic, expert-curated parser that required at least one explicit mention of “prominent epicardial adipose tissue noted” or “prominent epicardial fat [depot]”. This label was intentionally designed to favor specificity over sensitivity, capturing overt clinical recognition of marked epicardial adiposity rather than a quantitative threshold susceptible to view-, angle-, and operator-dependent variation in linear measurements. Notably, for the purpose of model training, we labeled studies without such a mention as negative, acknowledging that absent documentation does not necessarily establish absent disease. This was however deemed to be the most feasible strategy to develop an echocardiographic biomarker for detecting prominent epicardial adiposity while maximizing specificity. Training, validation, and test splits were derived from the patient-level PanEcho splits, with no patient overlap (**Supplemental Methods**).^18^

### Model architecture and fine-tuning

We built PanAdipo on the publicly released PanEcho video foundation model previously developed by our group.^18^ As previously described, the PanEcho backbone integrates standard echocardiographic views (PLAX, parasternal short-axis [PSAX], apical 2-, 3-, 4-, and 5-chamber [A2C, A3C, A4C, and A5C], subcostal, and Doppler) within a shared spatial encoder and a temporal transformer, producing a 768-dimensional pooled embedding from a 16-frame clip at 224×224 pixel resolution. To this embedding, we attached a lightweight binary classification head consisting of a dropout layer (rate 0.3) followed by a single linear projection, and we fine-tuned the full network end-to-end. We trained the network in distributed data-parallel mode across six NVIDIA H100 GPUs with an effective batch size of 144 video clips. We set the initial learning rate to 10^−4^ for the head (10-fold lower for the backbone) and used the AdamW optimizer. To address the expected low prevalence of the target label (2.3% of development studies), we applied a positive-class weight of 15 directly within the binary cross-entropy loss. We trained for up to 50 epochs with early stopping when the validation area under the precision-recall curve (AUPRC) failed to improve for five consecutive epochs, and retained the best checkpoint for downstream evaluation. At inference time, we generated a single per-study PanAdipo score by averaging the model outputs across all eligible clips for a given study, providing a view-pooled score that does not require a specific acquisition view.^18^ The resulting study-level output (range: 0-1) was operationalized as a continuous biomarker (score) of the prominent epicardial fat depot phenotype, with higher values denoting greater model confidence in the presence of prominent epicardial adiposity by echocardiography. Therefore, in our downstream analyses we opted against using a single diagnostic threshold. We also recognize that, because of the positive-class weight, the model’s default output should not be treated as a calibrated probability.

### Internal and external testing cohorts

We evaluated the PanAdipo model in four testing cohorts whose composition was chosen to stress test the model along four complementary axes: temporal, modality, geographic, and freedom from confounding by indication (**Central Illustration**). For initial internal testing, we used a temporally distinct YNHHS TTE set of patients sampled from the second half of 2022 (n=4,588 individuals contributing 5,130 unique studies), thus probing temporal generalization within the same institution and acquisition protocol. In this set, the label was ascertained by applying the same parser to report text from the echocardiography reporting system and the institutional imaging data warehouse. Recognizing the lack of standardization in the reporting of prominent epicardial adiposity, and to evaluate the external transportability of the biomarker, we further deployed PanAdipo to a held-out cohort of patients within YNHHS who underwent coronary computed tomography angiography (CCTA) within 12 months of a TTE (see “Cross-sectional radiomic body and organ analysis”), as well as three additional cohorts. The first was cardiac-focused POCUS performed in YNHHS emergency departments (n=10,957 studies from unique individuals), which captures abbreviated, off-axis acquisitions by frontline clinicians. The second was a geographically distinct sample of adult TTE studies from MIMIC-IV (n=4,549 studies from unique individuals), a publicly available critical care and hospital-based dataset from Beth Israel Deaconess Medical Center (Boston, MA, USA).^20,21^ MIMIC-IV represents an entirely separate health system, vendor mix, and patient population, and tests geographic and institutional transportability.^20,21^ The third was research-grade adult TTE from Exam 6 of MESA (n=2,740 studies from unique individuals), a prospective community-based cohort enrolled across six United States field centers without regard to cardiovascular symptoms; Exam 6 was conducted between 2016 and 2018 in middle-aged to older adults with detailed cardiometabolic phenotyping.^22^ MESA is therefore relatively free of the confounding-by-indication that affects clinically referred cohorts. Of note, because expert reader-defined EAT labels were available only within the YNHHS TTE cohort, the held-out (temporal) YNHHS TTE set was used to assess discrimination of prominent EAT, whereas the external cohorts, which lacked such labels, were used to evaluate the clinical and prognostic associations of the PanAdipo score (transportability).

### Benchmarking against existing PanEcho outputs

To test whether the prominent epicardial fat depot phenotype could be inferred from existing deep learning-derived echocardiographic measurements, we evaluated the overlap between PanAdipo and the panel of PanEcho structural and functional outputs in the held-out YNHHS TTE testing set, as well as the original corresponding TTE report measurements. We further compared PanAdipo with multivariable logistic models of age, sex, and BMI and of PanEcho outputs, fitted in a development case-control sample and evaluated in the test set (**Supplemental Methods**).

### Explainability and cross-modal radiomic analyses

#### Saliency mapping

Gradient-weighted class activation maps (Grad-CAM++) were generated from the final convolutional block of the PanEcho ConvNeXt image encoder for high-confidence true-positive predictions.^23^ Per-frame maps were computed independently across each 16-frame clip, sharpened with an unsharp mask, and thresholded at a normalized activation of 0.25 for visualization. These Grad-CAMs are presented for illustrative purposes.

#### Cross-sectional radiomic body and organ analysis

To examine cross-modal correspondence between PanAdipo and body- and organ-level adiposity phenotypes derived from CCTA, we identified, within YNHHS, all individuals who had undergone a CCTA (between 2013 and 2025) within one year of a TTE, and who are included in the multicenter Oxford Risk Factors and Non-invasive Imaging (ORFAN) study (NCT05169333).^11^ We then identified wide field-of-view (FoV) series that extended sufficiently to allow cross-sectional assessment of subcutaneous, intermuscular, and pericardial adiposity in addition to the cardiac chambers. We processed each CCTA volume with the open-source Body and Organ Analysis (BOA) pipeline,^24,25^ performing fully automated multi-organ segmentation that quantifies the volume and attenuation of adipose depots and the volumes of the cardiac structures (**Supplemental Table 1**). The primary analysis included patients not used for model development, with one pair per patient (n=5,167). BOA defines EAT as adipose tissue (−190 to −30 HU) within the pericardium and paracardial fat as adipose tissue in the mediastinum outside the pericardium. Because CT quantifies EAT independently of report documentation, it served as the quantitative reference standard; we report the AUROC for high EAT (EAT volume indexed to total heart volume ≥95th percentile within age-sex groups).^26^

### Cross-sectional cardiometabolic associations in MESA

In MESA, we evaluated the association between key cardiometabolic biomarkers (primary: log[homeostatic model assessment for insulin resistance (HOMA-IR)]; secondary/exploratory: triglycerides, high-density lipoprotein [HDL] and low-density lipoprotein [LDL] cholesterol, total cholesterol, fasting glucose, HbA1c, systolic and diastolic blood pressure, and estimated glomerular filtration rate using the CKD-EPI equation) and the standardized log(PanAdipo). We repeated all models in a normoglycemic subgroup (fasting glucose <100 mg/dL without treated diabetes; HbA1c and insulin were measured at a subset of field centers).

### Longitudinal cardiometabolic outcomes

In YNHHS (TTE and POCUS) and MIMIC-IV, where longitudinal follow-up was available, we evaluated the prognostic value of PanAdipo for four cardiometabolic endpoints (T2D, hypertension, hyperlipidemia, and metabolic dysfunction-associated steatotic liver disease and steatohepatitis [MASLD/MASH]), each defined from linked structured electronic health record (EHR) data as the first documentation after the index echocardiogram. Endpoints, and the prevalent disease excluded at baseline, were identified from structured diagnosis records using International Classification of Diseases, Ninth and Tenth Revision, Clinical Modification (ICD-9-CM and ICD-10-CM) codes (**Supplemental Table 2**). We further evaluated the association of PanAdipo with all-cause mortality in the same three cohorts and in MESA. To mitigate reverse causation due to prevalent disease, we restricted each analysis to individuals free of the respective condition at baseline and adopted a landmark design with a 90-day blanking window, as well as sensitivity analyses for 30 and 365 days. Deaths were treated as censoring events (cause-specific hazards), with Fine-Gray subdistribution models as a sensitivity analysis, and prevalent disease was defined using all available history.

### Statistical analysis

We summarized continuous variables as mean (standard deviation [SD]) or median [25^th^ to 75^th^ percentile] depending on distribution, and categorical variables as counts (percentages). We computed the area under the receiver operating characteristic curve (AUROC) and the AUPRC of PanAdipo for the binary label of prominent epicardial fat depot, overall, by view, and for view combinations (**Supplemental Methods**), with 95% confidence interval (CI) derived from 2,000 patient-level bootstrap replicates. To quantify the overlap of PanAdipo with existing echocardiographic measurements, we computed study-level Spearman correlations of PanAdipo with each feature available both as a report measurement and as a PanEcho output. For the paired CT analysis, we tested cross-modal associations between the per-study PanAdipo score and the CT adipose depot and cardiac structure measures (**Supplemental Table 1**) using Spearman rank correlations, with Holm-Bonferroni-adjusted P values; partial Spearman correlations were adjusted for subcutaneous adipose tissue (SAT) volume and BMI, additionally for EAT volume (paracardial adipose tissue), for EAT volume and tube voltage (attenuation), or for age, sex, and BMI.

In MESA, we modeled the cross-sectional association between log-transformed and standardized PanAdipo and each cardiometabolic biomarker (also z-scored) with ordinary least-squares linear regression, adjusting for age and sex, then additionally for BMI or, alternatively, waist-hip ratio (WHR), and, in a further model, for race and ethnicity, field center, and lipid-lowering, antihypertensive, and glucose-lowering medications, so that the coefficient on PanAdipo represents the standardized beta per SD of log(PanAdipo). Log HOMA-IR was designated the primary biomarker; other biomarkers were secondary or exploratory. Secondary and exploratory biomarkers were not adjusted for multiple comparisons and are interpreted descriptively.

For longitudinal analyses, we fit Cox proportional hazards models in each cohort separately with Huber-White robust standard errors, using log(PanAdipo), standardized within each cohort, as the exposure, with adjustment for age and sex in the primary model and additional adjustment for BMI in the secondary model. For the four cardiometabolic endpoints (T2D, hypertension, hyperlipidemia, MASLD/MASH), we pooled cohort-specific log hazard ratios using a random effects (restricted maximum likelihood) estimator with I^2^. Sensitivity analyses used Fine-Gray models with death as a competing event, age- and sex-adjusted models in the BMI-complete sample, and multiple imputation of missing BMI (**Supplemental Methods**). Tests were two-sided with a nominal significance level of 0.05 unless otherwise specified. All analyses were performed in Python 3.11 (package versions in the **Supplemental Methods**), and the analytical code is available at https://github.com/Cardiometabolic-Intelligence/PanAdipo-Manuscript-Analysis. Reporting follows the TRIPOD+AI statement,^27^ with a completed checklist provided in **Supplemental Table 3**.

## RESULTS

### Cohort composition

Across the four testing cohorts and the held-out CT-linked cohort, we analyzed 4,588 individuals in the YNHHS TTE held-out test set, 10,957 in YNHHS POCUS, 4,549 in MIMIC-IV, 2,740 in MESA, and 5,167 in the YNHHS TTE-CCTA cohort (**Table 1**, **Supplemental Figure 1**). The mean age ranged from 59±14 years (YNHHS TTE-CCTA; **Table 1**) to 74±9 years (MESA) (**Supplemental Figure 1A**), whereas the mean BMI ranged from 28.5 to 29.7 kg/m² (**Supplemental Figure 1B**); and PanAdipo scores followed right-skewed distributions across cohorts (median 0.05 to 0.09; IQR 0.03 to 0.18), with the lowest values in MESA (**Supplemental Figure 1C**, log scale). There was a balanced sex distribution, with the proportion of female participants ranging from 46.8% to 52.8% (among participants with recorded sex) across cohorts (**Table 1**). As expected for their respective settings, the hospital-based cohorts had a substantial baseline burden of cardiometabolic comorbidities, whereas the community-based MESA cohort had a markedly lower burden of overt cardiovascular disease but a high prevalence of hypertension and hyperlipidemia (**Table 1**).

**Table 1.** Baseline characteristics across the four PanAdipo testing cohorts and the held-out YNHHS TTE-CCTA cohort.

|  | 1. YNHHS<br>TTE | 2. YNHHS<br>POCUS | 3. MIMIC-IV TTE | 4. MESA<br>TTE | 5. YNHHS<br>TTE-CCTA |
| --- | --- | --- | --- | --- | --- |
| Total Counts (n) | 4,588 | 10,957 | 4,549 | 2,740 | 5,167 |
| Age, mean ± SD (years) | 65.5 ± 16.7 | 66.1 ± 17.2 | 70.3 ± 13.2 | 73.7 ± 8.5 | 59.0 ± 13.8 |
| Female, n (%) | 2,299 (50.4%) | 5,107 (46.8%) | 2,337 (52.6%) | 1,446 (52.8%) | 2,694 (52.1%) |
| Race and ethnicity |  |  |  |  |  |
| *Non-Hispanic White* | 3,324 (72.4%) | 6,986 (63.8%) | 2,891 (63.6%) | 1,116 (40.7%) | 3,450 (66.8%) |
| *Non-Hispanic Black* | 547 (11.9%) | 2,322 (21.2%) | 739 (16.2%) | 678 (24.7%) | 713 (13.8%) |
| *Hispanic* | 448 (9.8%) | 1,221 (11.1%) | 172 (3.8%) | 586 (21.4%) | 634 (12.3%) |
| *Asian* | 108 (2.4%) | 194 (1.8%) | 113 (2.5%) | 360 (13.1%) | 131 (2.5%) |
| *Other/Unknown* | 161 (3.5%) | 234 (2.1%) | 634 (13.9%) | - | 239 (4.6%) |
| BMI, mean ± SD (kg/m<sup>2</sup>) | 29.1 ± 7.1 | 29.7 ± 7.7 | 29.4 ± 7.2 | 28.5 ± 5.7 | 29.2 ± 6.5 |
| PanAdipo, median [IQR] | 0.08 [0.03-0.18] | 0.07 [0.03-0.17] | 0.09 [0.05-0.16] | 0.05 [0.03-0.07] | 0.07 [0.03-0.15] |
| Hypertension, n (%) | 2,643 (57.6%) | 7,020 (64.1%) | 2,949 (64.8%) | 1,808 (66.0%) | 2,855 (55.3%) |
| Hyperlipidemia, n (%) | 2,238 (48.8%) | 4,812 (43.9%) | 2,422 (53.2%) | 1,714 (62.6%) | 2,692 (52.1%) |
| Type 2 diabetes, n (%) | 939 (20.5%) | 3,196 (29.2%) | 1,278 (28.1%) | 651 (23.8%) | 905 (17.5%) |
| MASLD/MASH, n (%) | 232 (5.1%) | 513 (4.7%) | 114 (2.5%) | - | 371 (7.2%) |
| Myocardial infarction, n (%) | 406 (8.8%) | 2,574 (23.5%) | 795 (17.5%) | 83 (3.0%) | 379 (7.3%) |
| Heart failure, n (%) | 1,077 (23.5%) | 4,323 (39.5%) | 1,257 (27.6%) | 64 (2.3%) | 974 (18.9%) |
| Chronic coronary syndrome, n (%) | 1,110 (24.2%) | 3,328 (30.4%) | 1,377 (30.3%) | 195 (7.1%) | 1,961 (38.0%) |
| Stroke, n (%) | 459 (10.0%) | 1,288 (11.8%) | 371 (8.2%) | 77 (2.8%) | 449 (8.7%) |
| Atrial fibrillation, n (%) | 675 (14.7%) | 1,918 (17.5%) | 805 (17.7%) | 187 (6.8%) | 469 (9.1%) |
| Peripheral vascular disease, n (%) | 142 (3.1%) | 582 (5.3%) | 282 (6.2%) | 32 (1.2%) | 138 (2.7%) |

The YNHHS TTE-CCTA cohort comprises patients not used for model training or validation who had a CCTA within one year of a TTE (one pair per patient; age at CT). Percentages for female sex use participants with recorded sex as the denominator. Dashes indicate data not available.
**Abbreviations:** BMI: body mass index; CCTA: coronary computed tomography angiography; IQR: interquartile range; MASLD: metabolic dysfunction-associated steatotic liver disease; MASH: metabolic dysfunction-associated steatohepatitis; MESA: Multi-Ethnic Study of Atherosclerosis; POCUS: point-of-care ultrasound; SD: standard deviation; TTE: transthoracic echocardiography; YNHHS: Yale-New Haven Health System.

### PanAdipo detects prominent epicardial adiposity beyond individual structural measurements

In the temporally held-out YNHHS TTE test set, PanAdipo discriminated the expert reader’s prominent EAT with an AUROC of 0.86 (95% CI, 0.82-0.91) and an AUPRC of 0.13 (95% CI, 0.09-0.22) across all views (74 [1.4%] label-positive studies) (**Figure 1B**). View-specific AUROCs ranged from 0.79 to 0.90, with the highest discrimination in the A4C and PLAX views (0.90 and 0.89, respectively), whereas combining A4C and PLAX views yielded an AUROC of 0.93 (95% CI, 0.89-0.96) and an AUPRC of 0.22 (95% CI, 0.10-0.38) (**Figure 1B**). To assess whether this phenotype could already be inferred from existing PanEcho-derived measurements, we correlated PanAdipo with each measured and PanEcho-derived feature. Observed correlations were weak throughout (|ρ| ≤0.21 for measured and ≤0.29 for PanEcho-derived features; **Figure 1C**), the strongest measured correlates being interventricular septal and posterior wall thickness (ρ=0.21 and 0.20). Multivariable comparators fitted in the development set, including PanEcho outputs combined with age, sex, and BMI, discriminated less well than PanAdipo (AUROC 0.83 vs 0.88 in test studies with available BMI; see detailed results in **Supplemental Table 4**). Taken together, these findings suggest that prominent epicardial adiposity, although partially correlated with structural remodeling, is not fully captured by traditional echocardiographic models focusing on structural heart disease or echocardiography interpretation.

**Figure 1.**
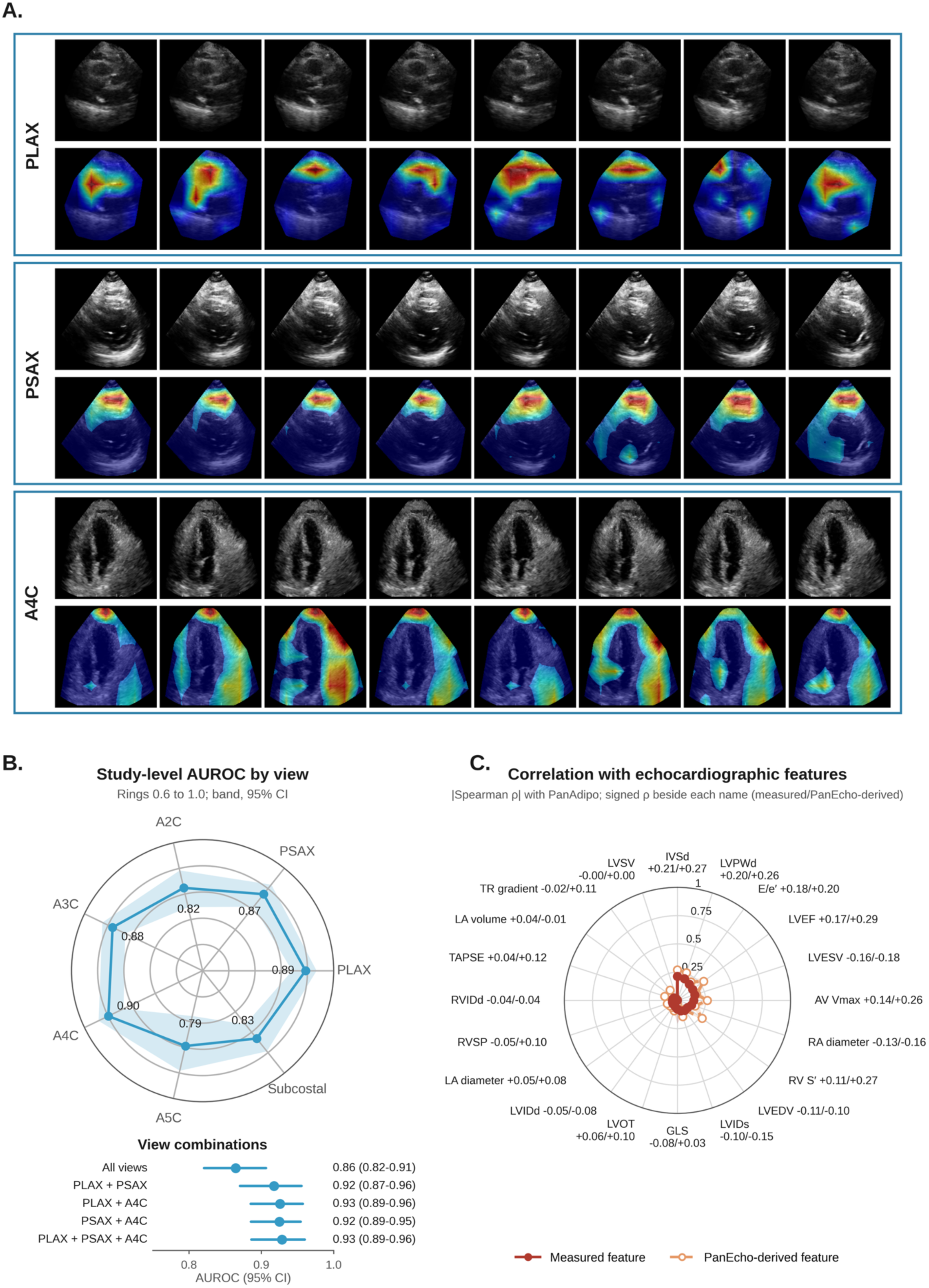
PanAdipo attends to the epicardial area, discriminates across views, and is only weakly correlated with conventional echocardiographic features. **(A**) Grad-CAM++ explainability maps from the PanAdipo video encoder, overlaid on representative frames across PLAX, PSAX, and A4C views. Attention largely concentrates over the epicardial area in each view. (B) Study-level AUROC (95% CI) for the report-derived label in the YNHHS TTE test set: top, by view (shaded band; rings 0.6 to 1.0; mean of that view’s standard on-axis clips within each study); bottom, all views pooled and combinations of on-axis PLAX, mid-papillary PSAX, and A4C views (mean of the view scores in studies with every view in the combination). (C) |Spearman ρ| of PanAdipo with each echocardiographic feature available both as a report measurement (solid) and as a PanEcho output (dashed) in the YNHHS TTE test set, with the signed ρ (measured/PanEcho-derived) beside each feature. **Abbreviations:** A4C, apical four-chamber; A2C, A3C, and A5C, apical 2-, 3-, and 5-chamber; Grad-CAM++, gradient-weighted class activation mapping; PLAX, parasternal long-axis; PSAX, parasternal short-axis; AV Vmax, aortic valve peak velocity; GLS, global longitudinal strain; IVSd, interventricular septum in diastole; LA, left atrial; LVEDV and LVESV, left ventricular end-diastolic and end-systolic volume; LVEF, left ventricular ejection fraction; LVIDd and LVIDs, left ventricular internal diameter in diastole and systole; LVOT, left ventricular outflow tract diameter; LVPWd, left ventricular posterior wall in diastole; LVSV, left ventricular stroke volume; RA, right atrial; RV S′, right ventricular systolic annular velocity; RVIDd, right ventricular internal diameter in diastole; RVSP, right ventricular systolic pressure; TAPSE, tricuspid annular plane systolic excursion; TR, tricuspid regurgitation.

### PanAdipo encodes specific radiomic phenotypes of epicardial adiposity

Grad-CAM++ saliency maps concentrated over the anterior epicardium in PLAX, over the anterior right ventricular wall in PSAX, and over the basal and apical epicardial regions in A4C views (**Figure 1A**; further false positive and negative examples are shown in **Supplemental Figure 2**). Because the model was weakly supervised, these maps indicate only that attention fell largely on the epicardial region rather than the cardiac chambers. As an independent test of the inferred phenotype and in an effort to provide a reader-independent ground truth label, we further identified individuals across YNHHS who had a CCTA within one year of the index echocardiogram (n=5,167 pairs from 5,167 unique participants; n=2,694 [52.1%] female participants, age 60 [IQR: 50-69] years) and computed the Spearman correlation between the per-study PanAdipo score and radiomic features extracted from automated body and organ segmentation of the CT scan (**Figure 2**). Among adipose and cardiac structure radiomic features, PanAdipo was most strongly associated with the indexed EAT volume to total heart volume (ρ=+0.76; 95% CI, 0.74-0.77; **Figures 2B** and **3A**). PanAdipo alone explained 53% of the variance in indexed EAT volume (R^2^ 0.53; 95% CI, 0.51-0.55), compared with 23% (0.21-0.25) for age, sex, and BMI combined, which added little beyond PanAdipo (R^2^ 0.54). Notably, the association of PanAdipo was attenuated for non-epicardial depots, such as subcutaneous adipose tissue volume (ρ=+0.45, P <0.001), or anthropometric indices such as BMI (ρ=+0.41, P <0.001). Moreover, the association with both absolute and indexed EAT volume persisted after adjustment for subcutaneous fat and BMI (partial ρ=0.60 for absolute and 0.70 for indexed EAT volume), whereas paracardial fat did not contribute beyond EAT (partial ρ=−0.02; **Supplemental Table 5**). Correlations with cardiac chamber, myocardial, and pericardial sac volumes were weak (|ρ| ≤0.19; **Figure 2B**), indicating that the score does not reflect cardiac size. Finally, PanAdipo’s correlation with indexed EAT volume (ρ=0.76) was broadly similar across demographic, acquisition, and pericardial effusion subgroups and for pairs acquired within 30 days (ρ=0.67-0.83; **Figure 3B**; **Supplemental Table 6**; **Supplemental Figure 3**). PanAdipo discriminated age- and sex-specific high EAT with AUROCs of 0.91-0.95 within age-sex groups, higher than in the pooled sample (AUROC 0.90; 95% CI, 0.89-0.92), underscoring the importance of age- and sex-specific evaluation and interpretation of the score (representative age- and sex-specific thresholds are presented in **Supplemental Table 7**).

**Figure 2.**
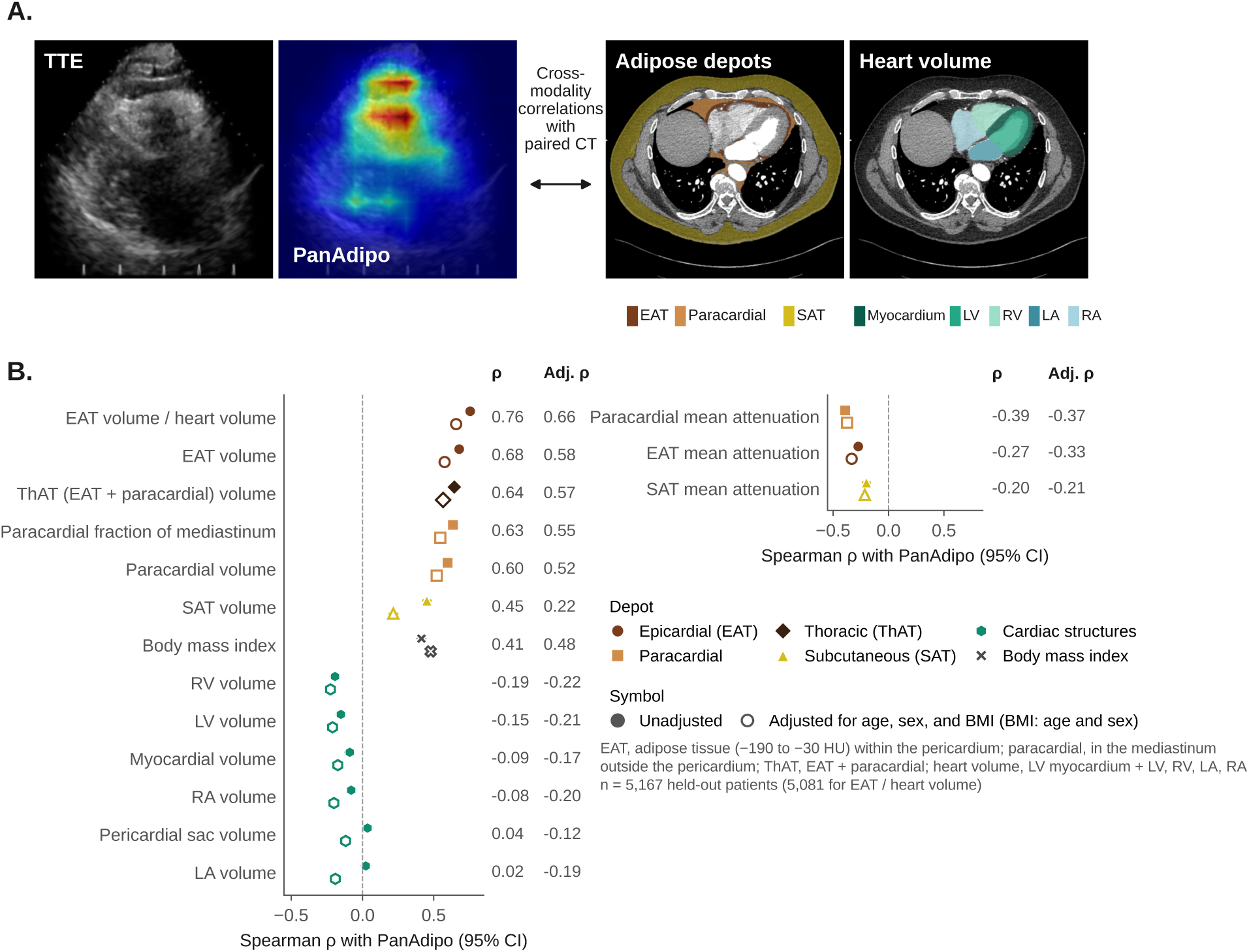
PanAdipo correlates with CT-derived epicardial adiposity. **(A)** Schematic of the cross-modal analysis in a TTE cohort at YNHHS with CCTA performed within one year of the index echocardiogram (n=5,167 pairs). The wide field-of-view of cardiac-focused CCTA allowed cross-sectional inclusion of epicardial, subcutaneous, paracardial, and total thoracic adiposity in addition to the cardiac chambers. PanAdipo scores from the echocardiogram were correlated with adipose depot and cardiac structure measures from automated body and organ analysis (BOA pipeline). Panels, left to right: mean parasternal long-axis frame and mean Grad-CAM++ map of high-confidence test clips, and an axial CCTA slice from one held-out patient with BOA segmentation of EAT, paracardial, and subcutaneous adipose tissue and of the cardiac structures summed as heart volume (LV myocardium, LV, RV, LA, and RA). (**B**) Spearman ρ (95% CI) of PanAdipo with adipose depot and cardiac structure volumes, EAT volume indexed to heart volume (sum of LV myocardium, LV, RV, LA, and RA volumes), the paracardial fraction of the mediastinum, and BMI (left) and mean depot attenuation (right); EAT, adipose tissue within the pericardium; paracardial, adipose tissue in the mediastinum outside the pericardium; ThAT, thoracic adipose tissue, their sum; rows are ordered by the magnitude of ρ; filled symbols, unadjusted; open symbols, adjusted for age, sex, and BMI (BMI itself for age and sex); data from 5,167 patients held out from model development (5,081 for EAT volume indexed to heart volume). **Abbreviations:** BOA, body and organ analysis; CCTA, coronary computed tomography angiography; CT, computed tomography; EAT, epicardial adipose tissue; TTE, transthoracic echocardiography; YNHHS, Yale-New Haven Health System.

**Figure 3.**
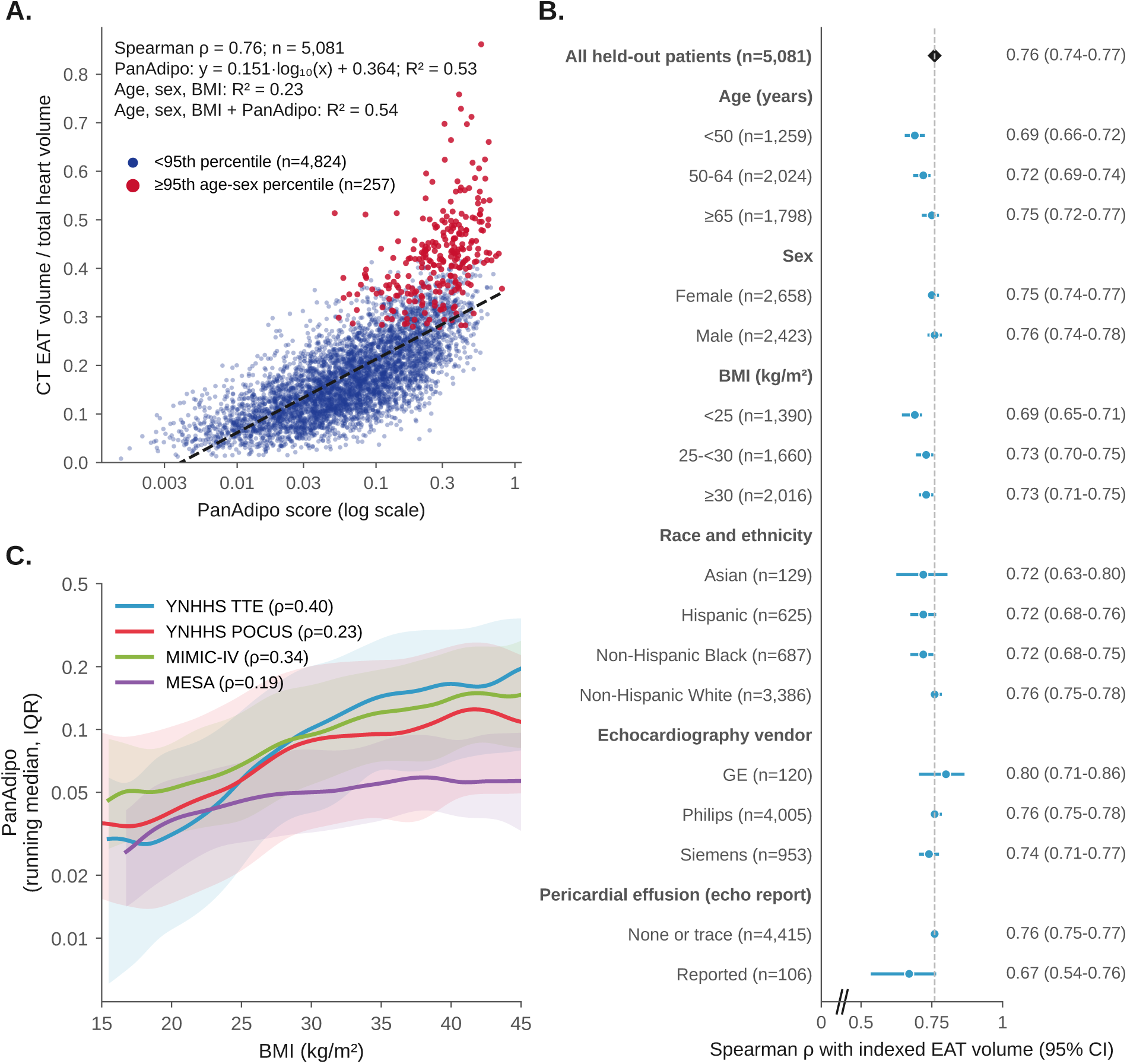
PanAdipo tracks heart volume-indexed epicardial fat consistently across subgroups and is only modestly correlated with body mass index. **(A)** CT EAT volume indexed to total heart volume against PanAdipo (log scale) in 5,081 patients held out from model development; red, at or above the 95th age- and sex-specific percentile (high EAT); navy, below; dashed line, least-squares fit of indexed EAT volume on log_10_(PanAdipo), with its formula and R^²^; R^²^ is also shown for a linear model of age, sex, and BMI without and with log_10_(PanAdipo) (5,066 patients with BMI). (B) Spearman ρ (95% CI) of PanAdipo with indexed EAT volume by age, sex, BMI, race and ethnicity, echocardiography vendor, and pericardial effusion reported on the echocardiogram; the dashed line marks the overall estimate; race and ethnicity other or unknown is not shown. (**C**) PanAdipo (log scale) against BMI in the YNHHS TTE test set, YNHHS POCUS, MIMIC-IV, and MESA; lines show the smoothed running median and shaded bands the IQR for BMI 15 to 45 kg/m², with Spearman correlation coefficients (ρ) per cohort (BMI 15 to 60 kg/m²). **Abbreviations:** BMI, body mass index; CT, computed tomography; EAT, epicardial adipose tissue; IQR, interquartile range; MESA, Multi-Ethnic Study of Atherosclerosis; POCUS, point-of-care ultrasound; TTE, transthoracic echocardiogram.

### Cross-sectional cardiometabolic associations in MESA persist after adjustment for BMI

Across cohorts, PanAdipo was only modestly correlated with BMI (Spearman ρ=0.40 in YNHHS TTE test, 0.34 in MIMIC-IV, 0.23 in YNHHS POCUS, and 0.19 in MESA; **Figure 3C**). To understand the BMI-independent implications of PanAdipo phenotyping, we evaluated cross-sectional associations with cardiometabolic biomarkers in the deeply phenotyped MESA cohort. Per SD of log(PanAdipo), there were significant associations with higher log(HOMA-IR), the primary biomarker, and with higher triglycerides and lower HDL cholesterol, and modestly higher diastolic blood pressure, with little or no association with LDL or total cholesterol (**Figure 4A**). These associations persisted after adjustment for BMI or WHR, after further adjustment for race and ethnicity, field center, and medications, and among normoglycemic participants (**Figure 4**; **Supplemental Table 8**); associations with triglycerides and HDL cholesterol further persisted among participants with BMI <25 kg/m² (**Supplemental Table 8**). Systolic blood pressure was not associated after BMI or WHR adjustment.

**Figure 4.**
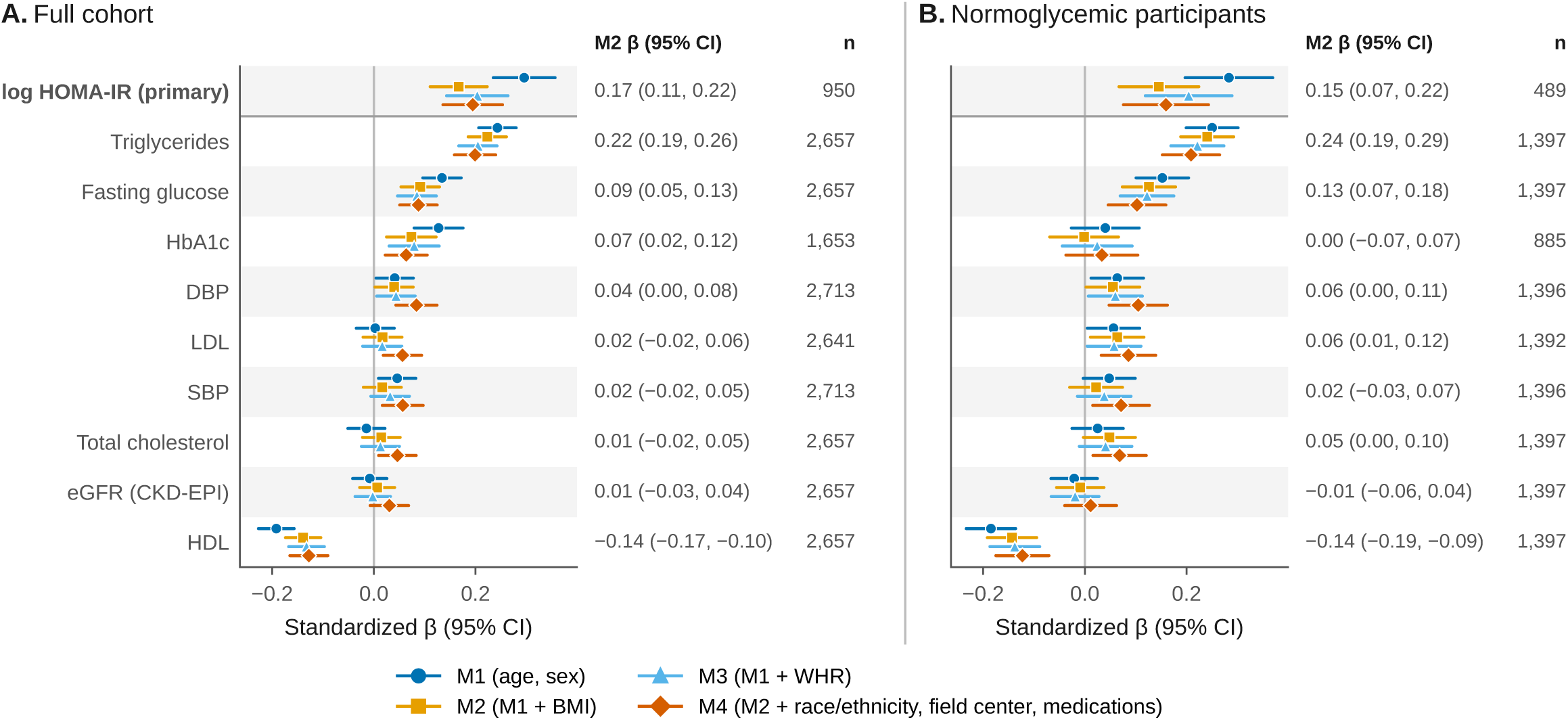
Cross-sectional cardiometabolic associations in MESA. Standardized beta coefficients (with 95% CIs) of log(PanAdipo) for cardiometabolic biomarkers measured at MESA Exam 6, in (**A**) the full cohort and (**B**) the normoglycemic subgroup (fasting glucose <100 mg/dL without treated diabetes). Models adjust for age and sex (M1), plus BMI (M2) or WHR (M3; an alternative to M2), and M2 plus race and ethnicity, field center, and medications (M4). Log HOMA-IR, the primary biomarker, is shown first; the other biomarkers are ordered by the BMI-adjusted (M2) estimate in the full cohort; values at right are M2 estimates, and n is the age- and sex-adjusted (M1) sample. **Abbreviations:** BMI, body mass index; DBP, diastolic blood pressure; eGFR, estimated glomerular filtration rate; HDL, high-density lipoprotein cholesterol; HOMA-IR, homeostatic model assessment for insulin resistance; LDL, low-density lipoprotein cholesterol; SBP, systolic blood pressure; WHR, waist-hip ratio.

### PanAdipo is associated with newly documented cardiometabolic disease in health systems

To evaluate the prognostic implications of opportunistic PanAdipo deployment, we performed landmark analyses across YNHHS (TTE and POCUS) and MIMIC-IV. In age- and sex-adjusted models, each SD of log(PanAdipo) was associated with higher risk of newly documented T2D, hyperlipidemia, and MASLD/MASH (90-day random-effects pooled HRs 1.28 [1.02-1.60], 1.17 [1.11-1.24], and 1.42 [1.18-1.69], respectively), but not of hypertension (1.13 [0.97-1.31]; **Figure 5**, **Supplemental Table 9**). Estimates for T2D and hypertension were heterogeneous across these differing cohorts, possibly reflecting differences in care setting (outpatient vs inpatient, elective vs urgent) (I²=81% for each; **Supplemental Table 10**). Prevalent disease excluded 27%, 63%, and 47% of participants for T2D, hypertension, and hyperlipidemia but only 4% for MASLD/MASH (**Supplemental Figure 4**), and Fine-Gray estimates accounting for the competing risk of death were similar (**Supplemental Table 10**). After additional adjustment for BMI in the subset with available anthropometrics, the associations with hyperlipidemia (HR 1.11, 95% CI 1.04-1.18) and MASLD/MASH (HR 1.31, 95% CI 1.03-1.67) persisted, including with multiple imputation of missing BMI and in Fine-Gray models, whereas those with T2D and hypertension did not (**Supplemental Table 10**). By contrast, mortality analyses showed an inverse association in the emergency department POCUS cohort, consistent with an obesity paradox pattern or reverse causation,^28^ but no association in the YNHHS TTE, MIMIC-IV, or community-based MESA cohorts (**Supplemental Table 11**).

**Figure 5.**
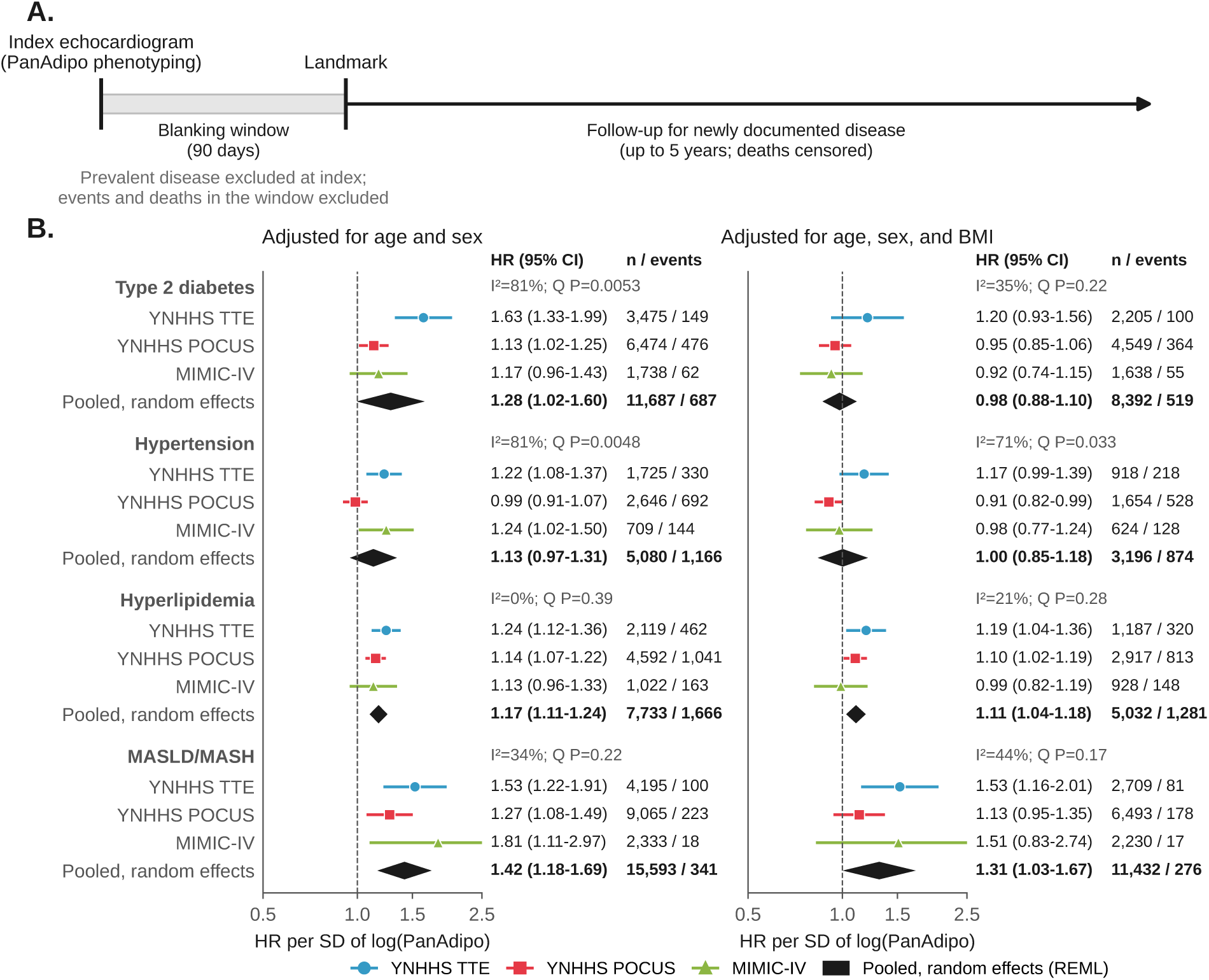
PanAdipo and newly documented metabolic disease in cohorts free of baseline disease. **(A)** *Landmark design*: individuals with baseline (prevalent) disease were excluded; follow-up for new-onset disease began after a 90-day blanking window following the index echocardiogram (30 and 365 days in sensitivity analyses), and individuals with the outcome or death during the window were excluded. **(B)** Forest plot of cohort-specific and pooled (random effects meta-analysis) hazard ratios per SD increase in log(PanAdipo) for newly documented type 2 diabetes, hypertension, hyperlipidemia, and MASLD/MASH at 90 days, adjusted for age and sex (left) and additionally for BMI (right), in the YNHHS (TTE and POCUS) and MIMIC-IV cohorts. Hazard ratios (95% CI), numbers of participants and events, and I^2^ with the Cochran Q P value are shown at right. **Abbreviations:** MASLD, metabolic dysfunction-associated steatotic liver disease; MASH, metabolic dysfunction-associated steatohepatitis; MIMIC, Medical Information Mart for Intensive Care; POCUS, point-of-care ultrasound; TTE, transthoracic echocardiogram.

## DISCUSSION

In this study, we developed PanAdipo, a video-based deep learning model that derives a phenotype of prominent epicardial adiposity from routine echocardiographic acquisitions, and evaluated its transportability and clinical relevance across diverse cohorts. We show that PanAdipo (i) identifies expert reader-defined prominent epicardial adiposity in a temporally held-out test cohort; (ii) captures information only partly represented by traditional anthropometric or echocardiographic measures of cardiac structure and function; (iii) discriminates and correlates most strongly with CT-defined epicardial adiposity; and (iv) is associated with insulin resistance, dyslipidemia, and subsequently documented MASLD/MASH and hyperlipidemia beyond conventional anthropometric measures.

Our findings carry several implications for the early recognition and management of CKM syndrome.^1^ First, echocardiography is the most widely performed cardiac imaging modality, and every standard transthoracic acquisition incidentally captures the epicardial fat compartment.^15,16^ PanAdipo enables scalable, opportunistic identification of prominent epicardial adiposity from imaging already obtained for unrelated indications, including abbreviated point-of-care studies performed by frontline clinicians, thus supporting both retrospective screening of existing echocardiographic archives and prospective phenotyping at the bedside via POCUS.^29^ Second, PanAdipo is only partly explained by anthropometric body size. It correlates only modestly with BMI and explains more than twice as much variance in CT-measured indexed EAT volume as age, sex, and BMI combined; its cross-sectional associations with HOMA-IR, triglycerides, and HDL persist after adjustment for anthropometrics; and its associations with newly documented MASLD/MASH and hyperlipidemia persist after adjustment for BMI. Third, if confirmed prospectively, identifying individuals with prominent epicardial adiposity who are missed by anthropometric risk assessment could inform the allocation of adipose-modifying therapies, including GLP-1 RA,^30^ dual or triple incretin agonists,^31,32^ SGLT2 inhibitors, and other CKM-targeted agents that act on the visceral and ectopic adipose milieu, and for trial enrichment in cardiometabolic prevention studies. Together, these properties suggest a complementary axis of CKM risk stratification, deliverable through imaging that is already in place.

Our observations sit naturally within the broader literature on epicardial and visceral adiposity. The prevalence of excess epicardial fat accumulation in unselected clinical populations is substantial,^8^ and EAT volume and composition have repeatedly been linked to insulin resistance and dyslipidemia independent of overall body size.^33,34^ The signature we observed, namely associations with HOMA-IR, triglycerides, and HDL cholesterol, but minimal associations with LDL cholesterol, total cholesterol, or systolic blood pressure, maps specifically to the insulin resistance and triglyceride-rich-lipoprotein axis of CKM, rather than to global, non-specific cardiovascular risk factors. These associations persist after adjustment for BMI or WHR and within a normoglycemic subgroup, supporting a signal not fully captured by anthropometric or glycemic measures alone.^8,35^ Linking prominent epicardial adiposity to downstream risk across the CKM spectrum, including atherosclerotic cardiovascular disease and heart failure with preserved ejection fraction, will require deeper phenotyping and longer follow-up.^36^ PanAdipo thus serves as a proof of concept that epicardial adiposity can be inferred from routine echocardiography.

### Study Limitations

Certain limitations merit consideration. First, the training label was an expert-reader assignment of a prominent epicardial fat depot extracted from routine clinical reports, which is more specific than sensitive. Report-negative studies were labeled negative, and external cohorts lacked report labels (MIMIC-IV echocardiography reports are not available); we therefore used CT-derived EAT as the quantitative reference standard. Clinical reports flag overt accumulation rather than applying a standardized echocardiographic threshold for epicardial adiposity. The continuous biomarker score nevertheless tracks a graded spectrum of severity, as supported by its correlation with CT-quantified EAT and its graded associations with cardiometabolic biomarkers and newly documented disease. Second, prognostic inference for hard cardiovascular outcomes is intrinsically difficult in clinically referred hospital cohorts. Patients undergoing TTE or POCUS are selected on cardiac symptoms or known disease, conditioning on the index echocardiogram introduces selection and collider bias, and competing risks and reverse causation through frailty and cachexia produce paradoxical short-term associations between adiposity and event rates.^28^ Our landmark analyses for newly documented T2D, hypertension, hyperlipidemia, and MASLD/MASH mitigate some of these biases, but prospective evaluation in lower-risk populations and over longer horizons will be needed to further clarify the prognostic value of this AI biomarker. Newly documented diagnoses may partly reflect surveillance and healthcare contact, although associations were consistent across cohorts and with MESA biomarkers, which are independent of healthcare contact; MASLD is underdiagnosed, and the attenuation of T2D and hypertension associations in BMI-adjusted models, which reflects both BMI adjustment and the smaller analytic sample, suggests that these associations are at least partly explained by body size. In addition, Asian participants, who carry higher cardiometabolic risk at lower BMI, were underrepresented, which may limit generalizability.^35,37^ Third, PanAdipo is not a replacement for cardiac CT-based radiomic profiling of EAT. With millimetric resolution and dedicated radiomic pipelines, CT captures quantitative and qualitative aspects of the epicardial fat compartment, including textural and density signatures that are not accessible to echocardiography.^5,11^ Finally, whether AI-derived phenotyping of prominent epicardial adiposity changes physician behavior, downstream investigation, and patient outcomes will require prospective, ideally randomized, evaluation.

## CONCLUSIONS

In conclusion, AI-enabled echocardiography provides a scalable biomarker of prominent epicardial adiposity that captures information beyond conventional echocardiographic and anthropometric measures, and is associated with an adverse cardiometabolic profile and subsequently documented metabolic disease. This approach may enable opportunistic metabolic risk stratification and provide a novel tool for clinical trials, including participant enrichment and longitudinal outcome assessment.

## DATA AND MODEL SHARING

Model weights and an inference code pipeline for PanAdipo are intended for release at the time of publication on GitHub (https://github.com/Cardiometabolic-Intelligence/PanAdipo), with an interactive dashboard accessible at: https://cardiometabolic-intelligence.github.io/PanAdipo. The analytical code used to generate the results, tables, and figures is available at https://github.com/Cardiometabolic-Intelligence/PanAdipo-Manuscript-Analysis. Underlying clinical imaging and electronic health record data from the Yale-New Haven Health System are not publicly redistributable. MIMIC-IV is available through PhysioNet credentialed access. This manuscript was prepared using MESA Research Materials obtained through the NHLBI Biologic Specimen and Data Repository Information Coordinating Center (BioLINCC) and/or the NHLBI BioData Catalyst^®^ ecosystem under approved data use agreements. The content is solely the responsibility of the authors and does not necessarily represent the official views of the MESA investigators, BioLINCC, BioData Catalyst, the NHLBI, or the National Institutes of Health (NIH).

## ACKNOWLEDGMENTS

The authors thank the participants and investigators of the Multi-Ethnic Study of Atherosclerosis (MESA), the MIMIC-IV team at Beth Israel Deaconess Medical Center, and the clinicians and staff of the Yale-New Haven Health System whose imaging studies made this work possible. The authors further wish to acknowledge the contributions of the consortium working on the development of the NHLBI BioData Catalyst® ecosystem, which enabled these analyses.

## PERSPECTIVES

### COMPETENCY IN MEDICAL KNOWLEDGE

A video-based deep learning model applied to standard transthoracic and point-of-care ultrasound acquisitions identifies prominent epicardial adiposity, captures information not represented by conventional echocardiographic measurements, corresponds most closely to CT-defined epicardial fat rather than to body mass index or other adipose compartments, and is associated, after adjustment for BMI, with insulin resistance, higher triglycerides, and subsequently documented metabolic disease.

### TRANSLATIONAL OUTLOOK

Prospective studies should determine whether point-of-care adiposity phenotyping refines metabolic risk assessment beyond anthropometry, whether the score tracks change with weight loss, incretin-based therapy, or lifestyle intervention, and whether it can enrich prevention trials with participants at increased cardiometabolic risk.

## ABBREVIATIONS AND ACRONYMS

BOA: Body and Organ Analysis
EAT: epicardial adipose tissue
FoV: field of view
Grad-CAM++: gradient-weighted class activation mapping
MESA: Multi-Ethnic Study of Atherosclerosis
MIMIC-IV: Medical Information Mart for Intensive Care, version IV
WHR: waist-hip ratio
YNHHS: Yale-New Haven Health System

---

**Central Illustration.**
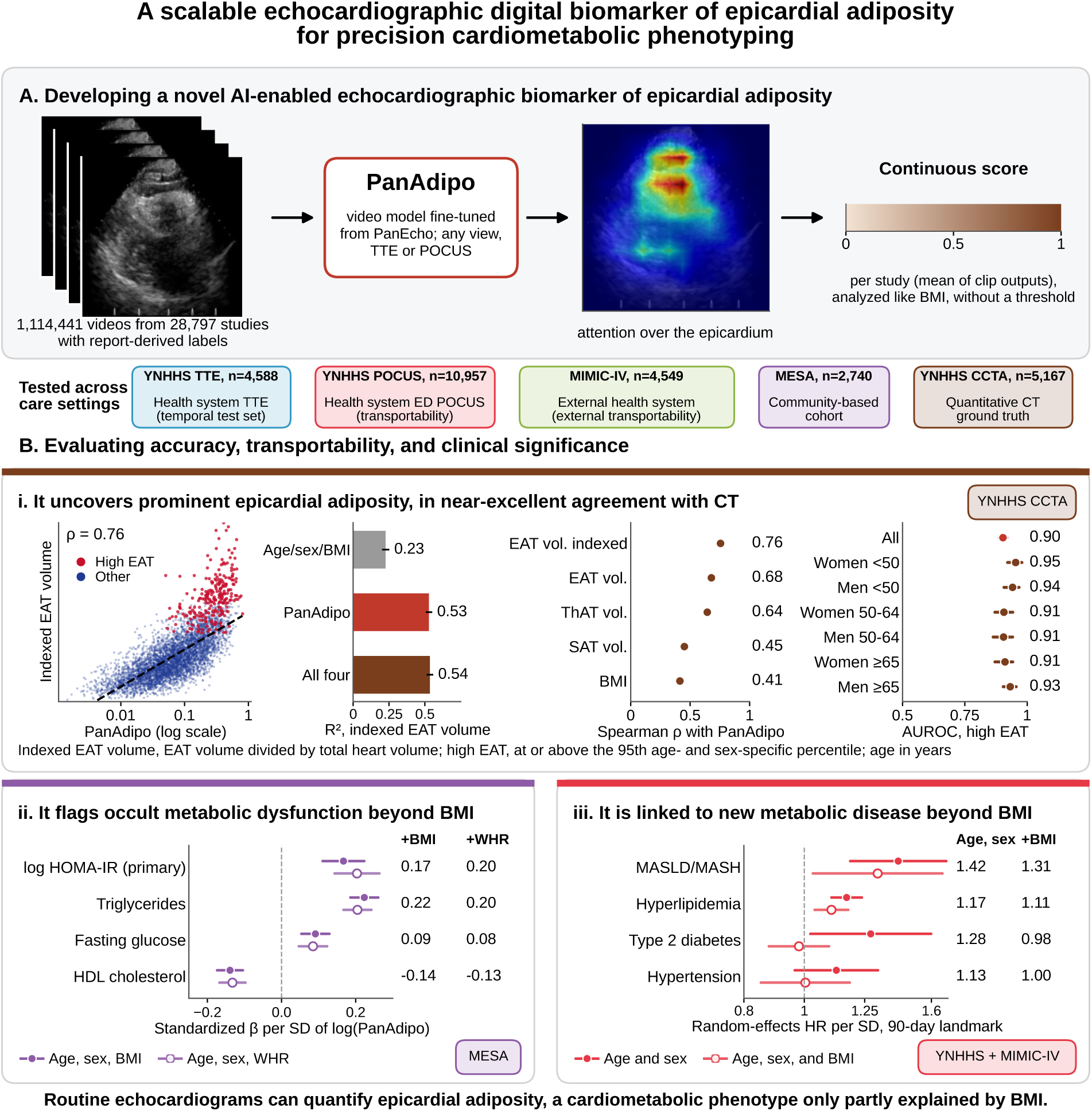
Study overview. **(A)** PanAdipo is a video-based deep learning model developed to characterize prominent epicardial adiposity (continuous score 0 to 1) from any standard 2D B-mode echocardiographic acquisition. The model was developed at the YNHHS and tested across four cohorts (a temporally distinct YNHHS TTE set, cardiac-focused emergency department POCUS, MIMIC-IV, and MESA) and in held-out YNHHS patients with paired CCTA. **(B)** Key findings: **i**, in held-out patients with paired CCTA, indexed EAT volume (EAT volume divided by total heart volume) against PanAdipo (log scale), with high EAT (at or above the 95th age- and sex-specific percentile) highlighted; variance in indexed EAT volume explained (R²) by age, sex, and BMI, by PanAdipo, and by all four; Spearman ρ of PanAdipo with CT adiposity measures and BMI; and AUROC for high EAT overall and by age-sex group; **ii**, standardized β per SD of log(PanAdipo) for cardiometabolic biomarkers in MESA, adjusted for age, sex, and BMI (filled) or for age, sex, and WHR (open); **iii**, random effects pooled hazard ratios per SD of log(PanAdipo) for newly documented metabolic disease at the 90-day landmark, adjusted for age and sex (filled) and additionally for BMI (open); all with 95% CIs. **Abbreviations:** AUROC, area under the receiver operating characteristic curve; BMI, body mass index; CCTA, coronary computed tomography angiography; CT, computed tomography; EAT, epicardial adipose tissue; ED, emergency department; HDL, high-density lipoprotein; HOMA-IR, homeostatic model assessment for insulin resistance; HR, hazard ratio; MASLD, metabolic dysfunction-associated steatotic liver disease; MASH, metabolic dysfunction-associated steatohepatitis; MESA: Multi-Ethnic Study of Atherosclerosis; MIMIC: Medical Information Mart for Intensive Care; POCUS: point-of-care ultrasound; SAT, subcutaneous adipose tissue; ThAT, thoracic adipose tissue; TTE, transthoracic echocardiography; WHR, waist-hip ratio; YNHHS, Yale-New Haven Health System.

