## Supplementary material for "AI-Enabled Echocardiography Identifies Prominent Epicardial Adipose Tissue Associated With an Adverse Cardiometabolic Profile": Online Supplement

**Supplemental Methods (addendum to main manuscript)**

***Label and partitions.*** The label was parsed from the Findings and Conclusions fields of the final report (case-insensitive phrase "prominent epicardial [fat/adipose]"); studies without the phrase were labeled negative. In the test set, the same rule was also applied to the report text (narrative and impression) archived in the institutional imaging data warehouse, and a study was labeled positive if either source matched (74 of 5,130 studies; 747 of 33,927 studies [2.2%] across the training, validation, and test sets). PanAdipo splits were derived from the patient-level PanEcho splits (training 19,868 patients, validation 1,611, test 4,588 patients contributing 5,130 studies; no patient overlap). In the CT cohort, 314 patients had contributed to training or validation and were excluded from the primary CT analysis; 324 patients appeared in both the YNHHS TTE and POCUS cohorts, and excluding them from POCUS changed pooled hazard ratios by at most 0.01.

***Model and score.*** The deployed model was fine-tuned on six GPUs with distributed data parallelism (24 clips per GPU; effective batch 144) using AdamW with learning rates of 1e-4 (head) and 1e-5 (backbone), weight decay 1e-4, dropout 0.3, a positive-class weight of 15 (no oversampling), mixed precision, and 16-frame clips from all views, with the learning rate reduced 10-fold after 3 epochs without improvement (minimum 1e-7), for up to 50 epochs with early stopping on validation AUPRC (patience 5). The study score is the mean of all clip outputs and is analyzed as a continuous score without a threshold. View-specific scores average the standard on-axis clips of each view; combination scores average the view scores of on-axis PLAX, mid-papillary PSAX, and A4C in studies with every view in the combination. CIs use 2,000 patient-level bootstrap replicates.

**Comparators.** To test whether existing measures jointly capture the phenotype, PanEcho (39 tasks; 40 outputs) was run on a development case-control sample (all 673 label-positive training and validation studies and 5,000 randomly selected label-negative studies; 224,555 videos). Logistic models on age, sex, and BMI, on the PanEcho study-level outputs (L2-penalized, penalty tuned by patient-grouped 5-fold cross-validation), and on both were fitted in this sample and evaluated in the test set; case-control sampling affects only the intercept (Supplemental Table 4).

**Label rates by reader.** In the development set, 76 of 133 reading cardiologists recorded the finding at least once. Among the 83 readers with at least 50 studies, the label rate ranged from 0% to 7.1% (median 2.2%; IQR, 0.9%-3.6%), and 13 never recorded it; the 10 readers with the most positive studies contributed 32% of positives from 21% of studies. Reader-level rates were only weakly consistent between the development and test periods (Spearman $\rho$=0.32 among 55 readers with at least 50 and 20 studies, respectively), consistent with reader-dependent reporting; this reader dependence motivated the use of CT-quantified EAT as a reader-independent reference standard.

***Grad-CAM++.*** PanAdipo is weakly supervised by study-level report labels, and Grad-CAM++ maps are only representative; they are used only to show that attention fell largely on the epicardial region rather than on the cardiac chambers (**Figure 1**). Representative false-positive and false-negative test studies are shown in Supplemental Figure 2.

***CT cohort.*** For each echocardiogram, the closest coronary computed tomography angiogram (CCTA) within 365 days was selected; the primary analysis used one pair per patient (shortest interval) among patients not used for model development (n=5,167). BOA defines adipose tissue as −190 to −30 HU; EAT (epicardial adipose tissue) lies within the pericardium and paracardial fat in the mediastinum outside the pericardium, so the fat fraction within the pericardium equals EAT volume divided by pericardial sac volume. Vendor, scanner, and tube voltage were extracted from DICOM headers. Because CT quantifies EAT independently of report documentation, it served as the quantitative reference standard. For the threshold analysis, high EAT was defined as EAT volume indexed to total heart volume at or above the 95th percentile within age (<50, 50-64, ≥65 years) and sex strata of the held-out sample. Total heart volume was the sum of the left ventricular myocardium, left ventricular cavity, right ventricle, right atrium, and left atrium including the appendage; segmentations with any structure <10 mL or a total outside 200-1,500 mL were excluded from this analysis.

***MESA and longitudinal analyses.*** Normoglycemia was defined by MESA variable dm036c (ADA 2003 classification, normal: fasting glucose <100 mg/dL without treated diabetes). HbA1c was not measured at two field centers and fasting insulin was measured in ancillary-study subsamples at four field centers; HOMA-IR was analyzed as a log-transformed, standardized variable, so estimates do not depend on the insulin assay unit, and absolute HOMA-IR values are not reported. In the hospital cohorts, time zero was the earliest echocardiogram; follow-up ended at the outcome, death (censored; cause-specific hazards), last recorded contact (MIMIC-IV), 5 years, or end of data; Fine-Gray models treated death before the endpoint as a competing event. Log(PanAdipo) was standardized within each cohort; standardization with a common SD changed pooled hazard ratios by at most 0.02. BMI was the measurement closest to the index study within ±365 days and was missing for 25%-35% of hospital-cohort participants (35.4% in YNHHS TTE, 30.3% in YNHHS POCUS, and 25.4% in MIMIC-IV). Participants without BMI had similar age (standardized difference −0.10 to 0.19), sex (absolute standardized difference ≤0.09), and PanAdipo scores (≤0.09) but fewer documented comorbidities, consistent with less contact with care: baseline hypertension was present in 43.9% vs 65.1% (YNHHS TTE), 53.1% vs 68.8% (YNHHS POCUS), and 43.7% vs 72.0% (MIMIC-IV) of those without vs with BMI (standardized differences 0.33-0.60), with similar differences for hyperlipidemia (0.40-0.50) and heart failure (0.19-0.53); in MIMIC-IV, race and ethnicity were more often unrecorded among those without BMI (32.5% vs 7.6%). Missing BMI was multiply imputed (20 imputations) by Bayesian linear regression of log BMI on age, sex, log(PanAdipo), the event indicator, the Nelson-Aalen cumulative hazard, and race and ethnicity within each cohort and endpoint landmark population; BMI-adjusted Cox models were combined with Rubin’s rules and pooled with random effects. In MIMIC-IV, diagnosis dates are admission dates and dates are shifted for deidentification; type 2 diabetes was defined from type-specific codes, and the 267 participants with any alcohol-related code were excluded from the MASLD/MASH analyses (Supplemental Table 2). Proportional-hazards (Schoenfeld) and log-linearity (spline) checks showed departures only for isolated cohort-endpoint combinations (diabetes in POCUS, P=0.008).

***Software.*** Python 3.11.14 (pandas 2.3.3, NumPy 2.3.3, SciPy 1.17.1, statsmodels 0.14.6, lifelines 0.30.3, scikit-learn 1.8.0); R 4.6.1 (survival 3.8.9) for Fine-Gray models; grad-cam 1.5.5 with

PyTorch 2.7.1. Analytical code is available at https://github.com/Cardiometabolic-Intelligence/PanAdipo-Manuscript-Analysis.

### Supplemental Table 1 | CT measures presented in the figures and tables.

| CT measure | Definition | Unit | Presented in |
|---|---|---|---|
| EAT volume indexed to total heart volume | EAT volume / total heart volume (LV myocardium + LV cavity + RV + RA + LA) | ratio | Central Illustration; Figures 2B, 3; Supplemental Tables 5-7 |
| Epicardial adipose tissue (EAT) volume | Adipose tissue (−190 to −30 HU) within the pericardium | mL | Central Illustration; Figure 2B; Supplemental Table 5 |
| EAT fraction of pericardial sac | EAT volume / pericardial sac volume | fraction | Supplemental Table 5 |
| Paracardial adipose tissue volume | Adipose tissue in the mediastinum outside the pericardium | mL | Figure 2B; Supplemental Table 5 |
| Paracardial fraction of mediastinum | Paracardial volume / mediastinal volume | fraction | Figure 2B |
| Thoracic adipose tissue (ThAT) volume | EAT + paracardial adipose tissue | mL | Figure 2B; Supplemental Table 5; Central Illustration |
| Subcutaneous adipose tissue (SAT) volume | Subcutaneous adipose tissue within the field of view | mL | Figure 2B; Supplemental Table 5; Central Illustration |
| Intermuscular adipose tissue volume | Adipose tissue between muscles within the field of view | mL | Supplemental Table 5 |
| Pericardial sac volume | Volume enclosed by the pericardium | mL | Figure 2B |
| LV, RV, LA, and RA volumes; myocardial volume | Cardiac chamber and LV myocardium segmentations | mL | Figure 2B; heart volume |
| Mean attenuation (EAT, paracardial, SAT) | Mean HU of the depot | HU | Figure 2B; Supplemental Table 5 (EAT) |
| EAT median attenuation | Median HU of EAT | HU | Supplemental Table 5 |

Measures from automated body and organ analysis (BOA) of coronary CT angiography; adipose tissue is defined as −190 to −30 HU.
**Abbreviations:** HU, Hounsfield units; LA, left atrium; LV, left ventricle; RA, right atrium; RV, right ventricle.

### Supplemental Table 2 | ICD diagnosis codes used to define newly documented cardiometabolic outcomes and prevalent disease.

| Outcome | YNHHS (ICD-10-CM) | MIMIC-IV, ICD-9-CM | MIMIC-IV, ICD-10-CM |
|---|---|---|---|
| Type 2 diabetes | E11.x; O24.1x | 250.x0, 250.x2 (x = 0-9; type 1 codes 250.x1 and 250.x3 not included) | E11.x; O24.1x |
| Hypertension | I10; I11.x; I12.x; I13.x; I67.4; O10.x; O11 | 401.x-405.x | I10; I11.x; I12.x; I13.x; I67.4; O10.x; O11 |
| Hyperlipidemia | E78.x | 272.0-272.4 | E78.x |
| MASLD/MASH | K76.0; K75.81 | 571.8† | K76.0; K75.81† |

The suffix ".x" denotes all subcodes within the listed three-character category.
†Participants with any alcohol-related code (ICD-9 291.x, 303.x, 571.0-571.3; ICD-10 F10.x, K70.x; n=267) were excluded.
Diagnoses were ascertained from structured electronic health record diagnosis tables by prefix matching on the listed codes, using all available history for prevalent disease. For each endpoint, the first qualifying diagnosis on or before the index echocardiogram defined prevalent disease and was excluded from the corresponding landmark analysis; the first qualifying diagnosis after the index echocardiogram defined the newly documented outcome.
**Abbreviations:** ICD-9-CM and ICD-10-CM, International Classification of Diseases, Ninth and Tenth Revisions, Clinical Modification; MASH, metabolic dysfunction-associated steatohepatitis; MASLD, metabolic dysfunction-associated steatotic liver disease; MIMIC: Medical Information Mart for Intensive Care; YNHHS, Yale-New Haven Health System.

### Supplemental Table 3 | Transparent Reporting of a multivariable prediction model for Individual Prognosis Or Diagnosis + Artificial Intelligence (TRIPOD+AI) checklist.

| Section / Topic | Item | Checklist item | Reported on page or section |
|---|---|---|---|
| **TITLE** | **1** | Identify the study as developing or evaluating the performance of a multivariable prediction model, the target population, and the outcome to be predicted | 1 |
| **ABSTRACT** | **2** | See TRIPOD+AI for Abstracts checklist | 2-4 |
| **INTRODUCTION** | **3a** | Healthcare context and rationale for developing the prediction model | 6-7 |
| | **3b** | Target population and intended purpose in the context of the care pathway | 6-7 |
| | **3c** | Known health inequalities between sociodemographic groups | NA |
| | **4** | Study objectives, including whether the study describes development or validation of a prediction model (or both) | 7 |
| **METHODS** | **5a** | Sources of data separately for the development and evaluation datasets, rationale, and representativeness | 8-11 |
| | **5b** | Dates of collected participant data, including start and end of accrual and end of follow-up | 8-11 |
| | **6a** | Study setting (e.g., primary, secondary, general population), number and location of centers | 8-11 |
| | **6b** | Eligibility criteria for study participants | 8-11 |
| | **6c** | Details of any treatments received and handling during model development or evaluation | 13-15 |
| | **7** | Data pre-processing and quality checking, including consistency across sociodemographic groups | 9-10; Supplemental Methods |
| | **8a** | Outcome being predicted, time horizon, rationale, and consistency of assessment | 8, 13-15 |
| | **8b** | If outcome assessment is subjective, qualifications and demographics of outcome assessors | NA |
| | **8c** | Any actions to blind assessment of the outcome to be predicted | NA |
| | **9a** | Choice of initial predictors and any pre-selection | 8-12 |
| | **9b** | Definition and timing of predictors, and any actions to blind assessment of predictors | 8-12 |
| | **9c** | Qualifications and demographics of predictor assessors | 8 |
| | **10** | How the study size was arrived at | 8-12 |
| | **11** | How missing data were handled | 13-15; Supplemental Methods; Supplemental Table 10 |
| | **12a** | How data were used for development and evaluation, including any partitioning | 8 |
| | **12b** | How predictors were handled (transformation, rescaling, standardization) | 9-10, 13-15 |
| | **12c** | Type of model, rationale, model-building steps, hyperparameter tuning, internal validation | 9-10 |
| | **12d** | If heterogeneity in performance was handled or quantified across clusters | 13-15 |
| | **12e** | Measures and plots used to evaluate model performance (discrimination, calibration, utility) | 13-15 |
| | **12f** | Any model updating arising from evaluation | N/A |
| | **12g** | For model evaluation, how predictions were calculated (formula, code, object, API) | 9-10 |
| | **13** | If class imbalance methods were used, justification and any recalibration | 9-10 |
| | **14** | Approaches to address model fairness and rationale | 17-18; Supplemental Table 6 |
| | **15** | Output of the prediction model and rationale for classification thresholds | 9-10 |
| | **16** | Differences between development and evaluation data (setting, eligibility, outcome, predictors) | 10-11 |
| | **17** | Institutional review board or ethics committee, consent or waiver | 7 |

| | | | |
|---|---|---|---|
| **OPEN SCIENCE** | **18a** | Source of funding and role of funders | 24 |
| | **18b** | Conflicts of interest and financial disclosures for all authors | 25- |
| | **18c** | Where the study protocol can be accessed or whether one was prepared | NA |
| | **18d** | Registration information for the study | Not registered |
| | **18e** | Availability of the study data | 23 |
| | **18f** | Availability of the analytical code | 23 |
| **PATIENT & PUBLIC INVOLVEMENT** | **19** | Patient and public involvement during design, conduct, reporting, interpretation, or dissemination | Not involved |
| **RESULTS** | **20a** | Participant flow, including with and without the outcome and follow-up time | 15, 19-20; Table 1; Supplemental Figure 4; Supplemental Table 9 |
| | **20b** | Characteristics for each data source / setting, including key dates, demographics, sample size, outcome events, follow-up, missingness | 15, Table 1, Supplement |
| | **20c** | For model evaluation, comparison with development data on distribution of important predictors | 15, Table 1, Supplement |
| | **21** | Number of participants and outcome events in each analysis | 15-20, Supplement |
| | **22** | Full prediction model details (formula, code, object, API) and access restrictions | 23 |
| | **23a** | Model performance estimates with confidence intervals, including subgroups | 15-20, Supplement |
| | **23b** | Heterogeneity of model performance across clusters, where examined | 15-20, Supplement |
| | **24** | Results of any model updating | N/A |
| **DISCUSSION** | **25** | Overall interpretation of main results, including fairness | 20-23 |
| | **26** | Limitations and their effects on biases, statistical uncertainty, generalizability | 20-23 |
| | **27a** | Assessment and handling of poor-quality or unavailable input data when implementing the model | 20-23 |
| | **27b** | Required user interaction with input data and required expertise | 20-23 |
| | **27c** | Next steps for future research with a view to applicability and generalizability | 20-23 |

### Supplemental Table 4 | Discrimination of the report label by PanAdipo and by multivariable comparators fitted in the development set, in the YNHHS TTE test set.

| Test sample | Model | Studies (positive) | AUROC (95% CI) | AUPRC (95% CI) | PanAdipo minus model, AUROC | PanAdipo minus model, AUPRC |
|---|---|---|---|---|---|---|
| All test studies | PanAdipo | 5,130 (74) | 0.86 (0.82-0.91) | 0.13 (0.09-0.22) | reference | reference |
| All test studies | PanEcho outputs (n=40) (L2 logistic) | 5,130 (74) | 0.80 (0.75-0.85) | 0.08 (0.05-0.13) | +0.07 (+0.04 to +0.09) | +0.06 (+0.01 to +0.11) |
| BMI-complete test studies | PanAdipo | 3,361 (51) | 0.88 (0.84-0.92) | 0.15 (0.09-0.27) | reference | reference |
| BMI-complete test studies | Age, sex, and BMI (logistic) | 3,361 (51) | 0.72 (0.65-0.79) | 0.04 (0.02-0.06) | +0.16 (+0.09 to +0.24) | +0.12 (+0.05 to +0.22) |
| BMI-complete test studies | PanEcho outputs (n=40) (L2 logistic) | 3,361 (51) | 0.80 (0.75-0.86) | 0.09 (0.05-0.17) | +0.08 (+0.05 to +0.11) | +0.06 (+0.01 to +0.14) |
| BMI-complete test studies | PanEcho outputs + age, sex, and BMI (L2 logistic) | 3,361 (51) | 0.83 (0.78-0.88) | 0.10 (0.06-0.18) | +0.05 (+0.03 to +0.08) | +0.05 (+0.00 to +0.13) |

Comparators were fitted on a development case-control sample (all 673 label-positive training and validation studies and 5,000 randomly selected label-negative studies; PanEcho outputs averaged over all 224,555 videos) and evaluated in the YNHHS TTE test set against the report label (74 positive studies); PanAdipo was not refitted. Models with BMI used the test studies with BMI. CIs and differences are from 2,000 paired patient-level bootstrap replicates.

**Abbreviations:** AUPRC, area under the precision-recall curve; AUROC, area under the receiver operating characteristic curve; BMI, body mass index; CI, confidence interval.

**Supplemental Table 5 | Correlation of PanAdipo with CT adipose depots, attenuation, and BMI in patients held out from model development.**

| CT measure | Patients | Spearman ρ (95% CI) | Holm P | Partial ρ (95% CI) | Partial correlation adjusted for |
|---|---|---|---|---|---|
| EAT fraction of pericardial sac volume | 5,167 | 0.74 (0.73-0.76) | <0.001 | 0.69 (0.67-0.70) | SAT volume, BMI |
| EAT volume indexed to total heart volume | 5,081 | 0.76 (0.74-0.77) | <0.001 | 0.70 (0.69-0.72) | SAT volume, BMI |
| EAT volume | 5,162 | 0.68 (0.66-0.69) | <0.001 | 0.60 (0.58-0.62) | SAT volume, BMI |
| Paracardial adipose tissue volume | 5,166 | 0.60 (0.58-0.61) | <0.001 | -0.02 (-0.05 to 0.01) | EAT volume, SAT volume, BMI |
| Thoracic adipose tissue (ThAT; epicardial + paracardial) volume | 5,161 | 0.64 (0.63-0.66) | <0.001 | | |
| Subcutaneous adipose tissue volume | 5,166 | 0.45 (0.43-0.47) | <0.001 | | |
| Intermuscular adipose tissue volume | 5,167 | 0.50 (0.48-0.52) | <0.001 | | |
| EAT mean attenuation | 5,162 | -0.27 (-0.30 to -0.25) | <0.001 | -0.21 (-0.24 to -0.19) | EAT volume, tube voltage |
| EAT median attenuation | 5,162 | -0.36 (-0.38 to -0.34) | <0.001 | -0.23 (-0.26 to -0.20) | EAT volume, tube voltage |
| Body mass index | 5,151 | 0.41 (0.39-0.44) | <0.001 | 0.48 (0.46-0.50) | Age, sex |

One echocardiogram-CCTA pair per patient among patients not used for model development (n=5,167). EAT, adipose tissue (−190 to −30 HU) within the pericardium; paracardial, adipose tissue in the mediastinum outside the pericardium; ThAT, thoracic adipose tissue, their sum; heart volume, sum of left ventricular myocardium, left ventricular cavity, right ventricle, right atrium, and left atrium (n=5,081 with plausible segmentation). Partial Spearman correlation adjusts the ranks of both variables for the ranks of the covariates. CIs are from 2,000 patient-level bootstrap replicates. Holm P, P value adjusted for multiple comparisons across the panel of CT measures (Holm step-down method).

**Abbreviations:** BMI, body mass index; CI, confidence interval; EAT, epicardial adipose tissue; SAT, subcutaneous adipose tissue.

### Supplemental Table 6 | Correlation of PanAdipo with EAT volume indexed to total heart volume across strata.

| Stratum or analysis | Level | Patients | Spearman ρ (95% CI) |
|---|---|---|---|
| All | Primary held-out sample | 5,081 | 0.76 (0.74-0.77) |
| Age (years) | <50 | 1,259 | 0.69 (0.66-0.72) |
| Age (years) | 50-64 | 2,024 | 0.72 (0.69-0.74) |
| Age (years) | >=65 | 1,798 | 0.75 (0.72-0.77) |
| Sex | Female | 2,658 | 0.75 (0.74-0.77) |
| Sex | Male | 2,423 | 0.76 (0.74-0.78) |
| BMI (kg/m2) | <25 | 1,390 | 0.69 (0.65-0.71) |
| BMI (kg/m2) | 25-<30 | 1,660 | 0.73 (0.70-0.75) |
| BMI (kg/m2) | >=30 | 2,016 | 0.73 (0.71-0.75) |
| Race and ethnicity | Asian | 129 | 0.72 (0.63-0.80) |
| Race and ethnicity | Hispanic | 625 | 0.72 (0.68-0.76) |
| Race and ethnicity | Non-Hispanic Black | 687 | 0.72 (0.68-0.75) |
| Race and ethnicity | Non-Hispanic White | 3,386 | 0.76 (0.75-0.78) |
| Race and ethnicity | Other or unknown | 254 | 0.73 (0.66-0.78) |
| Echo vendor | GE | 120 | 0.80 (0.71-0.86) |
| Echo vendor | Philips | 4,005 | 0.76 (0.75-0.78) |
| Echo vendor | Siemens | 953 | 0.74 (0.71-0.77) |
| CT scanner model | LightSpeed VCT | 108 | 0.83 (0.76-0.87) |
| CT scanner model | SOMATOM Definition Edge | 254 | 0.80 (0.74-0.84) |
| CT scanner model | SOMATOM Force | 4,717 | 0.75 (0.74-0.77) |
| Tube voltage (kVp) | 70-90 | 1,456 | 0.71 (0.68-0.74) |
| Tube voltage (kVp) | 100-110 | 1,337 | 0.74 (0.72-0.77) |
| Tube voltage (kVp) | 120 | 1,901 | 0.75 (0.73-0.77) |
| Tube voltage (kVp) | 130-140 | 385 | 0.72 (0.66-0.76) |
| Echocardiogram-CCTA interval | ≤90 days | 3,754 | 0.76 (0.74-0.77) |
| Echocardiogram-CCTA interval | ≤30 days | 2,509 | 0.76 (0.74-0.78) |
| Pericardial effusion | Excluding echo studies with reported effusion | 4,975 | 0.76 (0.74-0.77) |
| Pericardial effusion | Restricted to echo studies reporting no or trace effusion | 4,415 | 0.76 (0.75-0.77) |
| Pericardial effusion | Echo studies with reported effusion | 106 | 0.67 (0.54-0.76) |

Primary held-out sample (one pair per patient) with plausible cardiac segmentation; heart volume, sum of left ventricular myocardium, left ventricular cavity, right ventricle, right atrium, and left atrium. Race and ethnicity groups with fewer than 50 patients were combined. Vendor and scanner were abstracted from DICOM headers. CIs from bootstrap resampling.

**Abbreviations:** BMI, body mass index; CCTA, coronary computed tomography angiography; CI, confidence interval.

### Supplemental Table 7 | Detection of age- and sex-specific high EAT (EAT volume indexed to total heart volume ≥95th percentile) with age- and sex-specific PanAdipo thresholds.

| Group | Patients (high EAT; %) | AUROC (95% CI) | Threshold (95% interval) | Sensitivity, % | Specificity, % | PPV, % | NPV, % |
|---|---|---|---|---|---|---|---|
| All held-out patients | 5,081 (257; 5.1%) | 0.90 (0.89-0.92) | - | - | - | - | - |
| Female, age <50 years | 571 (29; 5.1%) | 0.95 (0.92-0.98) | 0.094 (0.058-0.133) | 93 | 87 | 28 | 100 |
| Male, age <50 years | 688 (35; 5.1%) | 0.94 (0.91-0.97) | 0.120 (0.072-0.134) | 89 | 90 | 31 | 99 |
| Female, age 50-64 years | 1,036 (52; 5.0%) | 0.91 (0.87-0.94) | 0.213 (0.125-0.220) | 81 | 87 | 25 | 99 |
| Male, age 50-64 years | 988 (50; 5.1%) | 0.91 (0.86-0.94) | 0.118 (0.118-0.218) | 92 | 75 | 16 | 99 |
| Female, age ≥65 years | 1,051 (53; 5.0%) | 0.91 (0.87-0.94) | 0.295 (0.190-0.352) | 83 | 87 | 25 | 99 |
| Male, age ≥65 years | 747 (38; 5.1%) | 0.93 (0.90-0.96) | 0.219 (0.164-0.227) | 92 | 86 | 26 | 100 |

Primary held-out sample (one pair per patient) with plausible cardiac segmentation. Total heart volume, sum of left ventricular myocardium, left ventricular cavity, right ventricle, right atrium, and left atrium including the appendage; segmentations with any structure <10 mL or a total outside 200-1,500 mL were excluded. High EAT, EAT volume indexed to total heart volume at or above the 95th percentile within age (<50, 50-64, ≥65 years) and sex strata of this sample, adapted from Molnar et al. Discrimination was higher within each age-sex group than in the pooled sample, in which high EAT is defined separately by group and PanAdipo score distributions differ, supporting age- and sex-specific evaluation and interpretation of the score. Thresholds maximize the Youden index within each age-sex group; their 95% intervals are from bootstrap resampling. Thresholds were selected in this sample and are hypothesis-generating; they require external calibration before any clinical use. CIs are from bootstrap resampling.

**Abbreviations:** AUROC, area under the receiver operating characteristic curve; CI, confidence interval; NPV, negative predictive value; PPV, positive predictive value.

### Supplemental Table 8 | Cross-sectional associations of PanAdipo with cardiometabolic biomarkers in MESA after additional adjustment.

| Biomarker | Full cohort, M2 | Full cohort, M3 | Full cohort, M4 | Normoglycemic, M2 | BMI <25 kg/m², M2 |
|---|---|---|---|---|---|
| log HOMA-IR | 0.17 (0.11-0.22); n=950 | 0.20 (0.14-0.26); n=950 | 0.19 (0.14-0.25); n=948 | 0.15 (0.07-0.22); n=489 | 0.10 (-0.05 to 0.25); n=202 |
| Triglycerides | 0.22 (0.19-0.26); n=2,657 | 0.20 (0.17-0.24); n=2,657 | 0.20 (0.16-0.24); n=2,645 | 0.24 (0.19-0.29); n=1,397 | 0.22 (0.15-0.30); n=742 |
| HDL | -0.14 (-0.17 to -0.10); n=2,657 | -0.13 (-0.17 to -0.10); n=2,657 | -0.13 (-0.17 to -0.09); n=2,645 | -0.14 (-0.19 to -0.09); n=1,397 | -0.15 (-0.22 to -0.08); n=742 |
| LDL | 0.02 (-0.02 to 0.06); n=2,641 | 0.02 (-0.02 to 0.06); n=2,641 | 0.06 (0.02-0.09); n=2,630 | 0.06 (0.01-0.12); n=1,392 | 0.07 (0.00-0.14); n=741 |
| Total cholesterol | 0.01 (-0.02 to 0.05); n=2,657 | 0.01 (-0.02 to 0.05); n=2,657 | 0.05 (0.01-0.08); n=2,645 | 0.05 (0.00-0.10); n=1,397 | 0.04 (-0.04 to 0.11); n=742 |
| Fasting glucose | 0.09 (0.05-0.13); n=2,657 | 0.08 (0.05-0.12); n=2,657 | 0.09 (0.05-0.12); n=2,645 | 0.13 (0.07-0.18); n=1,397 | 0.14 (0.07-0.22); n=742 |
| HbA1c | 0.07 (0.02-0.12); n=1,653 | 0.08 (0.03-0.13); n=1,653 | 0.06 (0.02-0.10); n=1,645 | 0.00 (-0.07 to 0.07); n=885 | 0.08 (-0.03 to 0.19); n=362 |
| SBP | 0.02 (-0.02 to 0.05); n=2,713 | 0.03 (-0.01 to 0.07); n=2,713 | 0.06 (0.02-0.10); n=2,702 | 0.02 (-0.03 to 0.07); n=1,396 | 0.02 (-0.05 to 0.09); n=766 |
| DBP | 0.04 (0.00-0.08); n=2,713 | 0.04 (0.01-0.08); n=2,713 | 0.08 (0.04-0.12); n=2,702 | 0.06 (0.00-0.11); n=1,396 | 0.05 (-0.02 to 0.12); n=766 |
| eGFR (CKD-EPI) | 0.01 (-0.03 to 0.04); n=2,657 | 0.00 (-0.04 to 0.03); n=2,657 | 0.03 (-0.01 to 0.07); n=2,645 | -0.01 (-0.06 to 0.04); n=1,397 | 0.03 (-0.03 to 0.10); n=742 |

Standardized β per SD of log(PanAdipo) (95% CI); n per model. M2, age, sex, and BMI; M3, age, sex, and WHR; M4, M2 plus race and ethnicity, field center, and lipid-lowering, antihypertensive, and glucose-lowering medications. Normoglycemic, fasting glucose <100 mg/dL without treated diabetes (MESA dm036c). HbA1c was not measured at two field centers and fasting insulin was measured in ancillary-study subsamples at four field centers. Log HOMA-IR was designated the primary biomarker as the most direct available measure of insulin resistance; the remaining biomarkers are secondary or exploratory and were not adjusted for multiple comparisons.
**Abbreviations:** BMI, body mass index; DBP, diastolic blood pressure; eGFR, estimated glomerular filtration rate; HDL, high-density lipoprotein cholesterol; HOMA-IR, homeostatic model assessment for insulin resistance; LDL, low-density lipoprotein cholesterol; SBP, systolic blood pressure; WHR, waist-hip ratio

### Supplemental Table 9 | Pooled hazard ratios for newly documented cardiometabolic disease.

| Outcome | Landmark (d) | Adjustment | n | Events | HR (95% CI) |
|---|---|---|---|---|---|
| Type 2 diabetes | 30 | Age, sex | 12,252 | 799 | 1.22 (0.96-1.54) |
| | | Age, sex, BMI | 8,807 | 603 | 0.94 (0.87-1.03) |
| | 90 | Age, sex | 11,687 | 687 | 1.28 (1.02-1.60) |
| | | Age, sex, BMI | 8,392 | 519 | 0.98 (0.88-1.10) |
| | 365 | Age, sex | 10,280 | 464 | 1.37 (1.09-1.71) |
| | | Age, sex, BMI | 7,171 | 344 | 1.04 (0.93-1.17) |
| Hypertension | 30 | Age, sex | 5,500 | 1,427 | 1.13 (0.97-1.32) |
| | | Age, sex, BMI | 3,512 | 1,087 | 1.02 (0.84-1.25) |
| | 90 | Age, sex | 5,080 | 1,166 | 1.13 (0.97-1.31) |
| | | Age, sex, BMI | 3,196 | 874 | 1.00 (0.85-1.18) |
| | 365 | Age, sex | 4,197 | 641 | 1.12 (0.97-1.29) |
| | | Age, sex, BMI | 2,452 | 441 | 0.99 (0.77-1.29) |
| Hyperlipidemia | 30 | Age, sex | 8,350 | 1,992 | 1.16 (1.09-1.24) |
| | | Age, sex, BMI | 5,489 | 1,552 | 1.10 (0.99-1.23) |
| | 90 | Age, sex | 7,733 | 1,666 | 1.17 (1.11-1.24) |
| | | Age, sex, BMI | 5,032 | 1,281 | 1.11 (1.04-1.18) |
| | 365 | Age, sex | 6,432 | 1,018 | 1.19 (1.11-1.27) |
| | | Age, sex, BMI | 3,959 | 745 | 1.11 (1.03-1.21) |
| MASLD/MASH | 30 | Age, sex | 16,306 | 376 | 1.40 (1.23-1.59) |
| | | Age, sex, BMI | 11,954 | 302 | 1.28 (1.02-1.61) |
| | 90 | Age, sex | 15,593 | 341 | 1.42 (1.18-1.69) |
| | | Age, sex, BMI | 11,432 | 276 | 1.31 (1.03-1.67) |
| | 365 | Age, sex | 13,747 | 236 | 1.52 (1.13-2.04) |
| | | Age, sex, BMI | 9,835 | 192 | 1.45 (0.99-2.13) |

Random-effects (REML) pooled hazard ratios per 1-SD increments in log[PanAdipo]. Sensitivity estimates are in Supplemental Table 10.

### Supplemental Table 9 (continued) | Cohort-specific hazard ratios and random-effects weights at the 90-day landmark.

| Outcome | Cohort | Age, sex: n / events | HR (95% CI) | Weight | Age, sex, BMI: n / events | HR (95% CI) | Weight |
|---|---|---|---|---|---|---|---|
| Type 2 diabetes | YNHHS TTE | 3,475 / 149 | 1.63 (1.33-1.99) | 31.0% | 2,205 / 100 | 1.20 (0.93-1.56) | 16.6% |
| Type 2 diabetes | YNHHS POCUS | 6,474 / 476 | 1.13 (1.02-1.25) | 38.2% | 4,549 / 364 | 0.95 (0.85-1.06) | 61.6% |
| Type 2 diabetes | MIMIC-IV | 1,738 / 62 | 1.17 (0.96-1.43) | 30.8% | 1,638 / 55 | 0.92 (0.74-1.15) | 21.8% |
| Hypertension | YNHHS TTE | 1,725 / 330 | 1.22 (1.08-1.37) | 34.6% | 918 / 218 | 1.17 (0.99-1.39) | 32.4% |
| Hypertension | YNHHS POCUS | 2,646 / 692 | 0.99 (0.91-1.07) | 39.2% | 1,654 / 528 | 0.91 (0.82-0.99) | 42.8% |
| Hypertension | MIMIC-IV | 709 / 144 | 1.24 (1.02-1.50) | 26.1% | 624 / 128 | 0.98 (0.77-1.24) | 24.8% |
| Hyperlipidemia | YNHHS TTE | 2,119 / 462 | 1.24 (1.12-1.36) | 30.1% | 1,187 / 320 | 1.19 (1.04-1.36) | 20.8% |
| Hyperlipidemia | YNHHS POCUS | 4,592 / 1,041 | 1.14 (1.07-1.22) | 58.5% | 2,917 / 813 | 1.10 (1.02-1.19) | 68.1% |
| Hyperlipidemia | MIMIC-IV | 1,022 / 163 | 1.13 (0.96-1.33) | 11.4% | 928 / 148 | 0.99 (0.82-1.19) | 11.0% |
| MASLD/MASH | YNHHS TTE | 4,195 / 100 | 1.53 (1.22-1.91) | 37.1% | 2,709 / 81 | 1.53 (1.16-2.01) | 36.5% |
| MASLD/MASH | YNHHS POCUS | 9,065 / 223 | 1.27 (1.08-1.49) | 51.6% | 6,493 / 178 | 1.13 (0.95-1.35) | 50.3% |
| MASLD/MASH | MIMIC-IV | 2,333 / 18 | 1.81 (1.11-2.97) | 11.3% | 2,230 / 17 | 1.51 (0.83-2.74) | 13.1% |

Per SD of log(PanAdipo) (95% CI), cause-specific Cox models in each cohort; weight, share of the random-effects (restricted maximum likelihood) pooled estimate. Pooled estimates are in Supplemental Table 9.

**Abbreviations:** BMI: body mass index; CI, confidence interval; HR: hazard ratio; MASH: metabolic dysfunction-associated steatohepatitis; MASLD: metabolic dysfunction-associated steatotic liver disease; MIMIC: Medical Information Mart for Intensive Care; POCUS: point-of-care ultrasound; TTE: transthoracic echocardiogram; YNHHS, Yale-New Haven Health System.

### Supplemental Table 10 | Random-effects pooled hazard ratios at the 90-day landmark with competing-risk, BMI-complete, and multiple-imputation sensitivity analyses.

| Outcome | Adjustment | n / events / deaths | HR (95% CI) | I² (95% CI) | Fine-Gray HR | BMI-complete age/sex HR or BMI-imputed HR |
|---|---|---|---|---|---|---|
| Type 2 diabetes | Age, sex | 11,687 / 687 / 1,198 | **1.28 (1.02-1.60)** | 80.9% (30.0%-99.6%) | **1.29 (1.04-1.60)** | 1.15 (0.98-1.35) |
| Type 2 diabetes | Age, sex, BMI | 8,392 / 519 / 915 | 0.98 (0.88-1.10) | 34.7% (0.0%-98.9%) | 0.99 (0.91-1.09) | 1.09 (0.89-1.33) |
| Hypertension | Age, sex | 5,080 / 1,166 / 241 | 1.13 (0.97-1.31) | 81.3% (24.5%-99.4%) | 1.13 (0.98-1.30) | 1.13 (0.92-1.38) |
| Hypertension | Age, sex, BMI | 3,196 / 874 / 165 | 1.00 (0.85-1.18) | 70.7% (0.0%-99.1%) | 1.01 (0.86-1.18) | 1.01 (0.91-1.14) |
| Hyperlipidemia | Age, sex | 7,733 / 1,666 / 654 | **1.17 (1.11-1.24)** | 0.0% (0.0%-97.4%) | **1.18 (1.12-1.24)** | **1.16 (1.09-1.23)** |
| Hyperlipidemia | Age, sex, BMI | 5,032 / 1,281 / 433 | **1.11 (1.04-1.18)** | 21.2% (0.0%-98.8%) | **1.12 (1.05-1.19)** | **1.12 (1.03-1.21)** |
| MASLD/MASH | Age, sex | 15,593 / 341 / 2,094 | **1.42 (1.18-1.69)** | 34.4% (0.0%-98.7%) | **1.42 (1.22-1.67)** | **1.48 (1.13-1.92)** |
| MASLD/MASH | Age, sex, BMI | 11,432 / 276 / 1,598 | **1.31 (1.03-1.67)** | 44.4% (0.0%-98.0%) | **1.32 (1.06-1.64)** | **1.24 (1.09-1.41)** |

Per SD of log(PanAdipo) (95% CI), pooled across YNHHS TTE, YNHHS POCUS, and MIMIC-IV with random effects (restricted maximum likelihood); $\tau^2$ is imprecise with three cohorts. HRs are cause-specific (deaths censored). Fine-Gray, subdistribution HR with death before the endpoint as the competing event (deaths, competing deaths in the analytic sample). Last column: for age/sex rows, the age- and sex-adjusted HR re-estimated in the BMI-complete sample; for BMI rows, a sensitivity analysis in which missing BMI was multiply imputed (20 imputations) before BMI adjustment. Main BMI-adjusted estimates are complete-case. Imputation and pooling are described in the Supplemental Methods.

### Supplemental Table 10 (continued) | Incremental discrimination of PanAdipo for newly documented MASLD/MASH at the 90-day landmark.

| Cohort | Patients / events | C-statistic, age, sex, BMI | C-statistic, plus PanAdipo | Change in C-statistic (95% CI) |
|---|---|---|---|---|
| YNHHS TTE | 2,709 / 81 | 0.679 | 0.699 | +0.021 (0.000 to +0.059) |
| YNHHS POCUS | 6,493 / 178 | 0.641 | 0.644 | +0.003 (-0.003 to +0.022) |
| MIMIC-IV | 2,230 / 17 | 0.708 | 0.714 | +0.006 (-0.023 to +0.085) |

Harrell's C-statistic of Cox models with age, sex, and BMI, without and with standardized log(PanAdipo), in the BMI-complete 90-day landmark population of each cohort; apparent (in-sample) estimates. 95% CIs for the change are from 1,000 patient-level bootstrap replicates in which both models were refitted. This was a post hoc, exploratory analysis.

**Abbreviations:** BMI: body mass index; CI, confidence interval; HR: hazard ratio; MASH: metabolic dysfunction-associated steatohepatitis; MASLD: metabolic dysfunction-associated steatotic liver disease; MIMIC: Medical Information Mart for Intensive Care; POCUS: point-of-care ultrasound; TTE: transthoracic echocardiogram; YNHHS, Yale-New Haven Health System.

### Supplemental Table 11 | Per-cohort hazard ratios for all-cause mortality.

| Cohort | Landmark (d) | Adjustment | n | Events | HR (95% CI) |
|---|---|---|---|---|---|
| YNHHS TTE | 30 | Age, sex | 4,488 | 385 | 0.90 (0.81-1.01) |
| | | Age, sex, BMI | 2,929 | 268 | 0.99 (0.84-1.16) |
| | 90 | Age, sex | 4,439 | 336 | 0.91 (0.81-1.02) |
| | | Age, sex, BMI | 2,902 | 241 | 0.98 (0.83-1.17) |
| | 365 | Age, sex | 4,326 | 223 | 0.89 (0.77-1.03) |
| | | Age, sex, BMI | 2,829 | 168 | 0.93 (0.77-1.13) |
| YNHHS POCUS | 30 | Age, sex | 9,990 | 2,209 | 0.81 (0.78-0.85) |
| | | Age, sex, BMI | 7,188 | 1,639 | 0.83 (0.79-0.87) |
| | 90 | Age, sex | 9,602 | 1,821 | 0.82 (0.78-0.85) |
| | | Age, sex, BMI | 6,949 | 1,400 | 0.83 (0.79-0.87) |
| | 365 | Age, sex | 8,939 | 1,158 | 0.81 (0.77-0.86) |
| | | Age, sex, BMI | 6,469 | 920 | 0.83 (0.78-0.88) |
| MIMIC-IV | 30 | Age, sex | 2,900 | 146 | 0.87 (0.73-1.02) |
| | | Age, sex, BMI | 2,769 | 143 | 0.95 (0.79-1.14) |
| | 90 | Age, sex | 2,603 | 114 | 0.91 (0.76-1.10) |
| | | Age, sex, BMI | 2,492 | 111 | 1.01 (0.82-1.25) |
| | 365 | Age, sex | 1,471 | 52 | 0.95 (0.72-1.27) |
| | | Age, sex, BMI | 1,377 | 50 | 1.07 (0.78-1.46) |
| MESA | 30 | Age, sex | 2,719 | 124 | 0.89 (0.73-1.09) |
| | | Age, sex, BMI | 2,717 | 123 | 0.89 (0.73-1.10) |
| | 90 | Age, sex | 2,717 | 124 | 0.89 (0.73-1.09) |
| | | Age, sex, BMI | 2,715 | 123 | 0.89 (0.73-1.10) |
| | 365 | Age, sex | 2,687 | 108 | 0.95 (0.77-1.17) |
| | | Age, sex, BMI | 2,685 | 107 | 0.95 (0.76-1.18) |

Per 1-standard deviation increments in log[PanAdipo]
**Abbreviations:** BMI: body mass index; CI, confidence interval; HR: hazard ratio; MESA: Multi-Ethnic Study of Atherosclerosis; MIMIC: Medical Information Mart for Intensive Care; POCUS: point-of-care ultrasound; TTE: transthoracic echocardiogram; YNHHS, Yale-New Haven Health System.

**Supplemental Figure 1 | Study Population Characteristics.**

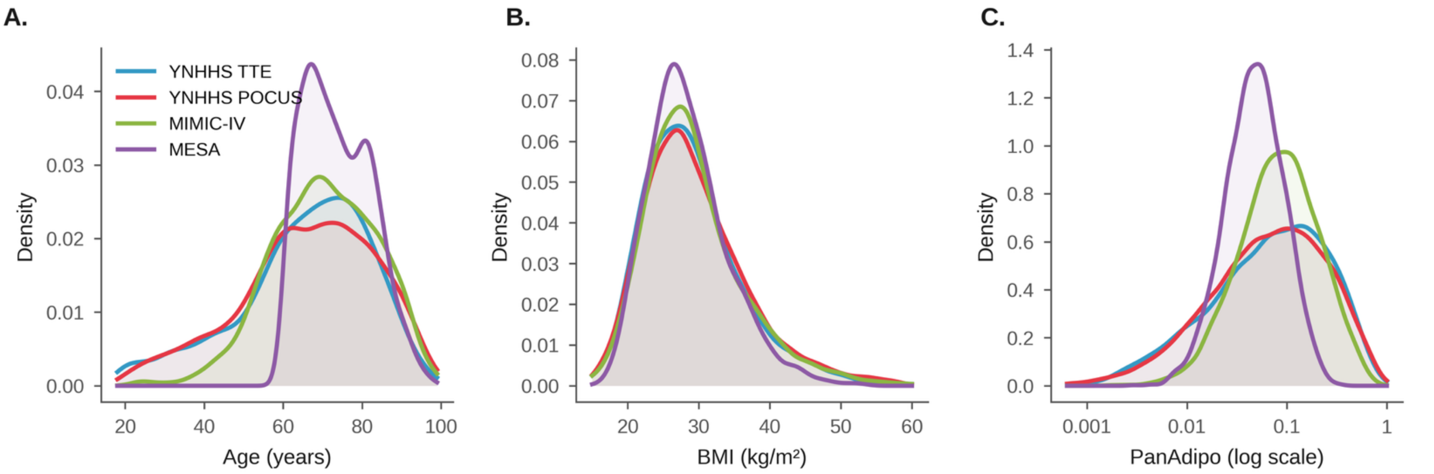

Cohort distributions of age (**A**), body mass index (**B**), and PanAdipo score (**C**; log scale) across the four PanAdipo testing cohorts (YNHHS TTE test, YNHHS POCUS, MIMIC-IV, and MESA); sex and baseline comorbidities are shown in Table 1.
**Abbreviations:** BMI, body mass index; MESA: Multi-Ethnic Study of Atherosclerosis; MIMIC: Medical Information Mart for Intensive Care; POCUS: point-of-care ultrasound; TTE: transthoracic echocardiogram; YNHHS, Yale-New Haven Health System.

**Supplemental Figure 2 | Grad-CAM++ maps in misclassified YNHHS TTE test studies.**

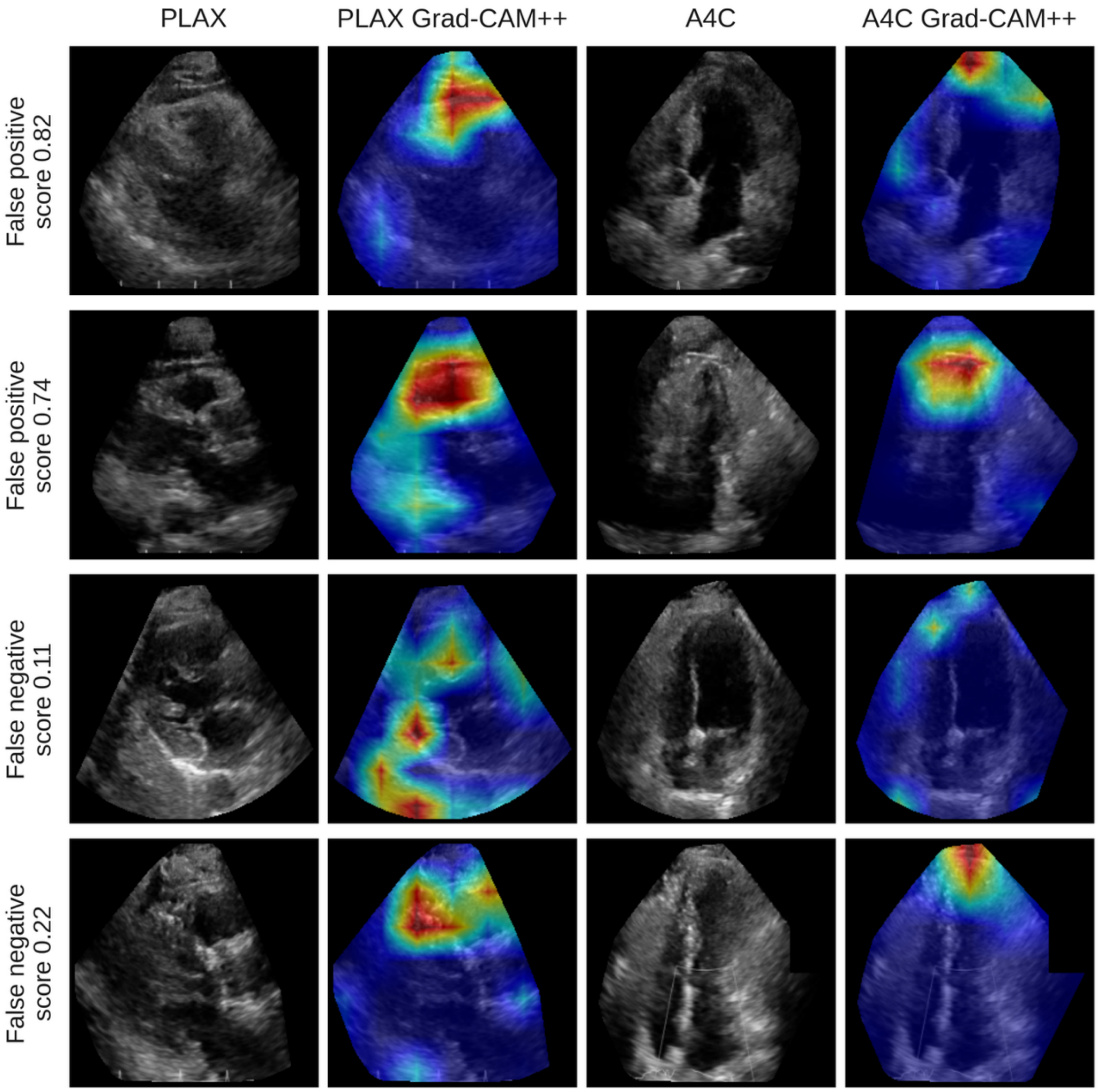

Frames from the first on-axis PLAX and A4C videos of two false-positive studies (report-negative; among the four highest PanAdipo scores in report-negative studies with report text) and two false-negative studies (report-positive; among the four lowest scores), against the report label. Cases were selected by rank before the maps were viewed. Maps were generated with Grad-CAM++ from the deployed model. Scores are study-level PanAdipo scores.

**Abbreviations:** A4C, apical 4-chamber; Grad-CAM++, gradient-weighted class activation mapping; PLAX, parasternal long axis; TTE: transthoracic echocardiogram; YNHHS: Yale-New Haven Health System.

**Supplemental Figure 3 | Interval between the echocardiogram and CCTA.**

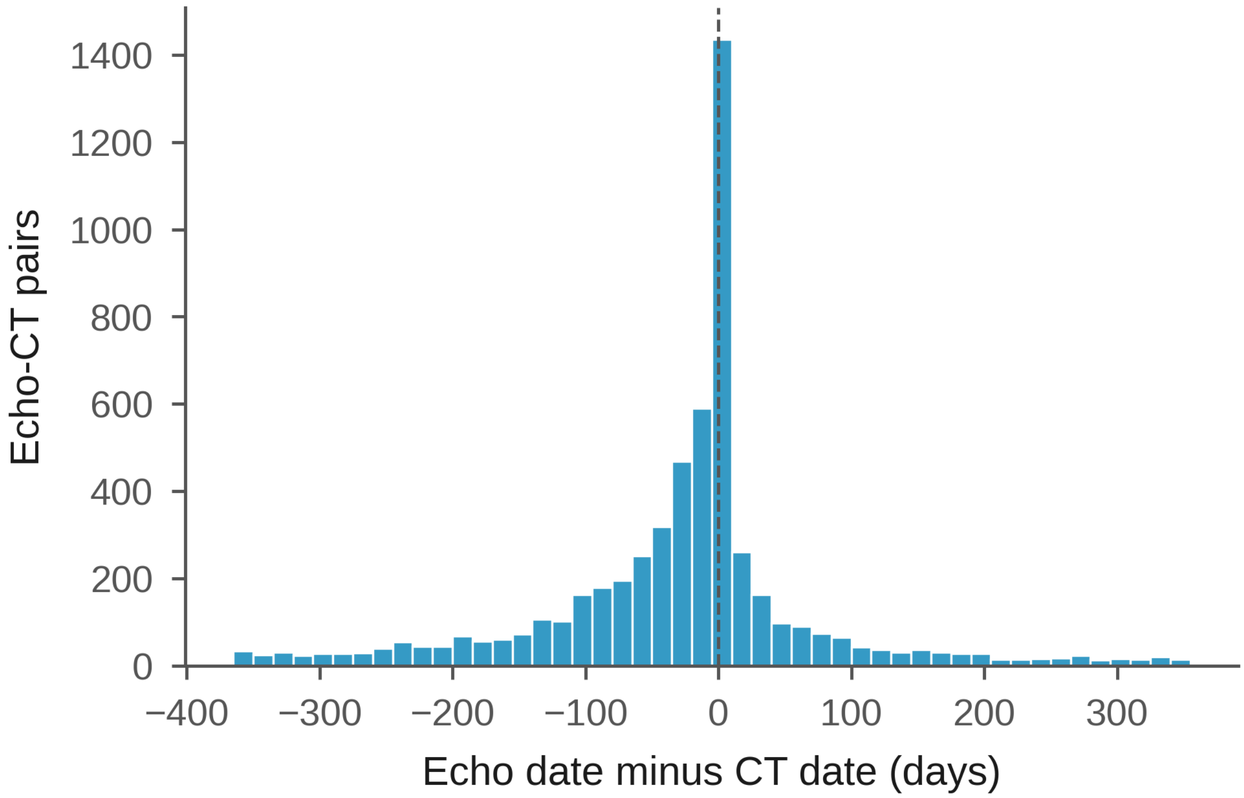

Distribution of the echocardiogram date minus the CCTA date for the echocardiogram-CCTA pairs (closest CCTA within 365 days).

### Supplemental Figure 4 | Participant flow for the 90-day landmark analyses.

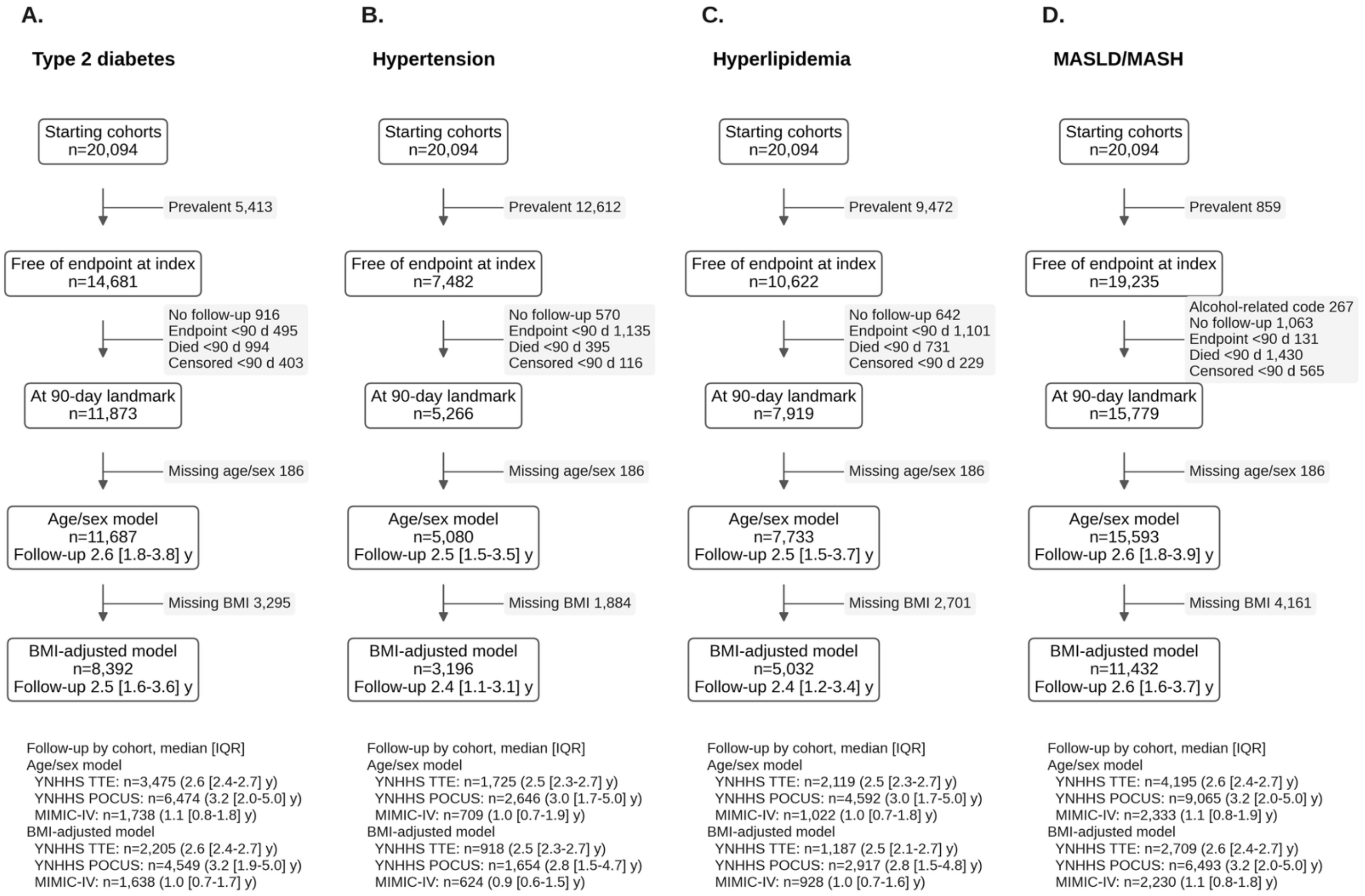

Exclusions from the starting cohorts (YNHHS TTE, YNHHS POCUS, and MIMIC-IV combined) to the age- and sex-adjusted and BMI-adjusted analytic samples for each endpoint; the analytic boxes give the median [IQR] follow-up from the index echocardiogram, in years, and the analytic n and follow-up of each cohort are listed below each panel.

**Abbreviations:** BMI: body mass index; IQR, interquartile range; MASH: metabolic dysfunction-associated steatohepatitis; MASLD: metabolic dysfunction-associated steatotic liver disease; MIMIC: Medical Information Mart for Intensive Care; POCUS: point-of-care ultrasound; TTE: transthoracic echocardiogram; YNHHS, Yale-New Haven Health System.
